# Neurofluid circulation changes during a focused attention style of mindfulness meditation

**DOI:** 10.1101/2025.09.10.25335492

**Authors:** Bryce A. Keating, David Vago, Kilian Hett, Ciaran Considine, Maria Garza, Caleb Han, Colin McKnight, Daniel O. Claassen, Manus J. Donahue

## Abstract

Neurofluids, including cerebrospinal fluid (CSF) and interstitial fluid, circulate through regulated central nervous system pathways to clear cerebral waste and support brain health, with elevated CSF flow hyperdynamicity and regurgitation through the cerebral aqueduct associating with aging and neurodegeneration. Sleep exerts state-dependent effects on neurofluid circulation, yet similar modulation during unique waking states, such as meditation, remains underexplored. Notably, mindfulness meditation shares several regulatory features with sleep, with core meditation practices representing distinct arousal states. We investigated whether the focused attention (FA) style of mindfulness meditation modulates neurofluid dynamics directionally opposite to aging and consistent with sleep. Using phase-contrast MRI, we assessed absolute CSF flow and velocity through the aqueduct, and using blood-oxygenation-level-dependent (BOLD) MRI, we assessed CSF fluctuations near the cervicomedullary junction together with total supratentorial gray matter fluctuations. Assessments were repeated in meditation-naïve adults during mind wandering (MW) without (n=13; repeatability controls) and with (n=14; breath controls) respiration rate modulation, and in adept meditators (n=23) during MW and FA meditation. No aqueduct CSF flow changes were observed in control groups. In meditators, absolute aqueduct absolute CSF flow decreased from MW to FA meditation (4.60±2.27 mL/min to 4.17±2.10 mL/min, p=0.005) owing to reduced regurgitant cranially-directed CSF velocity. On BOLD, this paralleled increased low frequency (0.0614-0.0887 Hz) CSF fluctuations (p=0.0138), which were inversely correlated with gray matter fluctuations during FA. Findings suggest that mindfulness meditation may represent a non-pharmacological, waking state capable of modulating neurofluid dynamics in a directionally similar manner to sleep and opposite to aging and neurodegeneration.

**Significance statement:** Mindfulness meditation is widely recognized for its self-reported mental and physical health benefits, yet its effects on core physiological systems that support brain health remain incompletely understood. This study provides the first evidence that a focused attention (FA) style of mindfulness meditation can modulate cerebrospinal fluid (CSF) dynamics in humans. Using neuroimaging, we demonstrate that FA meditation reduces regurgitant CSF flow through the aqueduct, directionally opposite to patterns seen in aging and neurodegeneration; additionally, meditation-induced CSF changes near the skull base are similar to those reported during sleep, an enhancer of neurofluid circulation. Findings suggest that mindfulness may offer a novel, non-pharmacological, waking model for augmenting neurofluid circulation and provide a potential physiological mechanism linking meditation to brain health.

## Introduction

Neurofluid, including cerebrospinal fluid (CSF) and interstitial fluid (ISF), circulation plays a fundamental role in clearing cerebral waste products, maintaining central nervous system homeostasis, and promoting overall brain health (1, 2). The majority of CSF is produced in the atria of the lateral ventricles via filtration of arterial blood through choroid plexus epithelia (3). Subsequently, CSF traverses the cerebral aqueduct, where it mixes with a smaller amount of CSF produced in the fourth ventricle, and enters the subarachnoid space through the Foramen of Magendie and Luschka, circulates through the subarachnoid space, and is resorbed into the vascular system via arachnoid granulations at the level of the dural sinuses (4). More recent studies suggest that neurofluids also traverse the cerebral parenchyma along periarterial, perivenous, and interstitial spaces, as part of the glial-lymphatic (i.e., glymphatic) system (1, 5). Impaired neurofluid circulation has been linked to aging, cognitive decline, and neurodegeneration (6, 7), yet, abilities to modulate neurofluid circulation either pharmacologically or behaviorally are limited.

Accumulating evidence suggests that neurofluid circulation is state-dependent and sensitive to changes in arousal, vascular tone, respiratory, and neuronal oscillatory activity (8–11). Sleep has been reported to exert a profound influence on neurofluid motion and metabolic clearance, suggesting that neurofluid circulation and associated cerebral waste clearance may account for some of the restorative functions provided by sleep (2, 7, 9). For example, net CSF flow velocity through the cerebral aqueduct has been shown to vary with circadian rhythm, reducing to approximately 30% of its nighttime maximum velocity at midday (12). Similarly, a 95% reduction in the concentration of injected CSF tracers observed via two-photon imaging in periarterial and parenchymal spaces was reported in wake versus sleeping mice, suggesting greater CSF motion during sleep (2). Quantitative and direct measures of CSF flow have also been assessed during wake and sleep in humans using hemodynamic and fluid inflow indices from blood oxygenation level-dependent (BOLD) MRI. Notably, CSF inflow signal assessed at the level of the fourth ventricle, a surrogate of CSF motion, was observed to increase during stable non-rapid eye movement (NREM) sleep and fluctuations were anti-correlated with total cortical hemodynamic signal fluctuations (9). Uniquely, the large amplitude pulsatility of the CSF signal fluctuations during sleep was found at low frequencies (0-0.1 Hz, peaking at 0.05 Hz) and was tightly entrained to slow delta wave power (1-4 Hz) on electroencephalography (EEG), suggesting neurofluid motion is at least partly modulated by neural oscillation (9). Quantitative and direct MRI measures of CSF flow in adults through the aqueduct have also recently demonstrated that total CSF motion over the cardiac cycle becomes more hyperdynamic (i.e., higher retrograde diastolic flow and higher anterograde systolic flow) with age (13) and neurodegeneration (14), and preliminary evidence has suggested that these phenomena correlate with cerebral peptide retention in the setting of neurodegenerative proteinopathies (15, 16). Collectively, these findings suggest that neurofluid circulation is highly state-dependent, deteriorates when those states become dysregulated with age or neurodegeneration, and can be investigated non-invasively *in vivo* with appropriately parameterized neuroimaging methods.

These findings also raise the possibility that other physiologically distinct states, such as meditation, may similarly modulate neurofluid dynamics and offer non-pharmacological routes to support brain health. In support of this possibility, sleep and mindfulness meditation share several similarities in physiology and health-promoting benefits, such as decreased metabolic activity, sympathetic nervous system activity, cardiac and respiratory frequencies, blood pressure, circulating norepinephrine and cortisol, proinflammatory processes, and conversely increased post-activity mental clarity, relaxation, and energy (17–20). Both sleep and mindfulness meditation have been demonstrated to involve significant functional and connectivity changes in thalamocortical, hippocampal-cortical, and default mode network (DMN) activity (21–25), coupled with similar changes in rhythmic brain activity from normal waking states, suggesting potential common mechanisms capable of facilitating neurofluid circulation (26). Given the similarities in restorative and regulatory functions across autonomic, neurophysiological, and metabolic systems between sleep and meditation, we hypothesized that mindfulness meditation may offer a unique, non-pharmacological waking model for probing, and perhaps augmenting, neurofluid circulation.

To evaluate this hypothesis, we applied multi-contrast MRI to assess absolute CSF flow at the level of the cerebral aqueduct in sequence with CSF motion at the approximate level of the skull base and cervicomedullary junction. We tested the primary hypothesis that in contrast to repeated assessments in healthy adults during mind wandering (MW) states, the absolute motion of CSF at the level of the aqueduct becomes less hyperdynamic with a focused attention (FA) style of mindfulness meditation performed by adept meditators, opposite to what occurs with increasing age and neurodegeneration (13, 14). A secondary hypothesis was that anticorrelations between low frequency total supratentorial gray matter and CSF signal fluctuations near the skull base would occur during FA, analogous to what has recently been reported during sleep (9). If confirmed, findings would provide a new potential physiological mechanism linking meditation to brain health.

## Results

### Participants

Participant demographic characteristics are summarized in **Table 1**. In total, 52 participants were recruited, comprised of 25 adept meditators and 27 controls (13 repeatability controls and 14 breath controls). Two meditators were excluded as one did not meet radiological inclusion criteria and one phase contrast scan was motion corrupted, which yielded a final cohort of 23 meditators; all repeatability and breath controls met inclusion criteria. Cohorts were matched for sex and mean age within one decade of life and control participants were meditation naïve with less than 25 hours of lifetime meditation experience. Conversely, the meditation cohort had 3,708±4,759 lifetime hours of formal meditation experience with a median lifetime meditation experience of 2,390 hours. **Figure 1A** summarizes the experimental paradigm with timing, whereas the relevant post-processing steps for the CSF phase contrast and BOLD assessments are shown in **Figure 1B** and **Figure 1C**, respectively, with additional technical details provided in **Supporting Information**.

**Figure 1.**
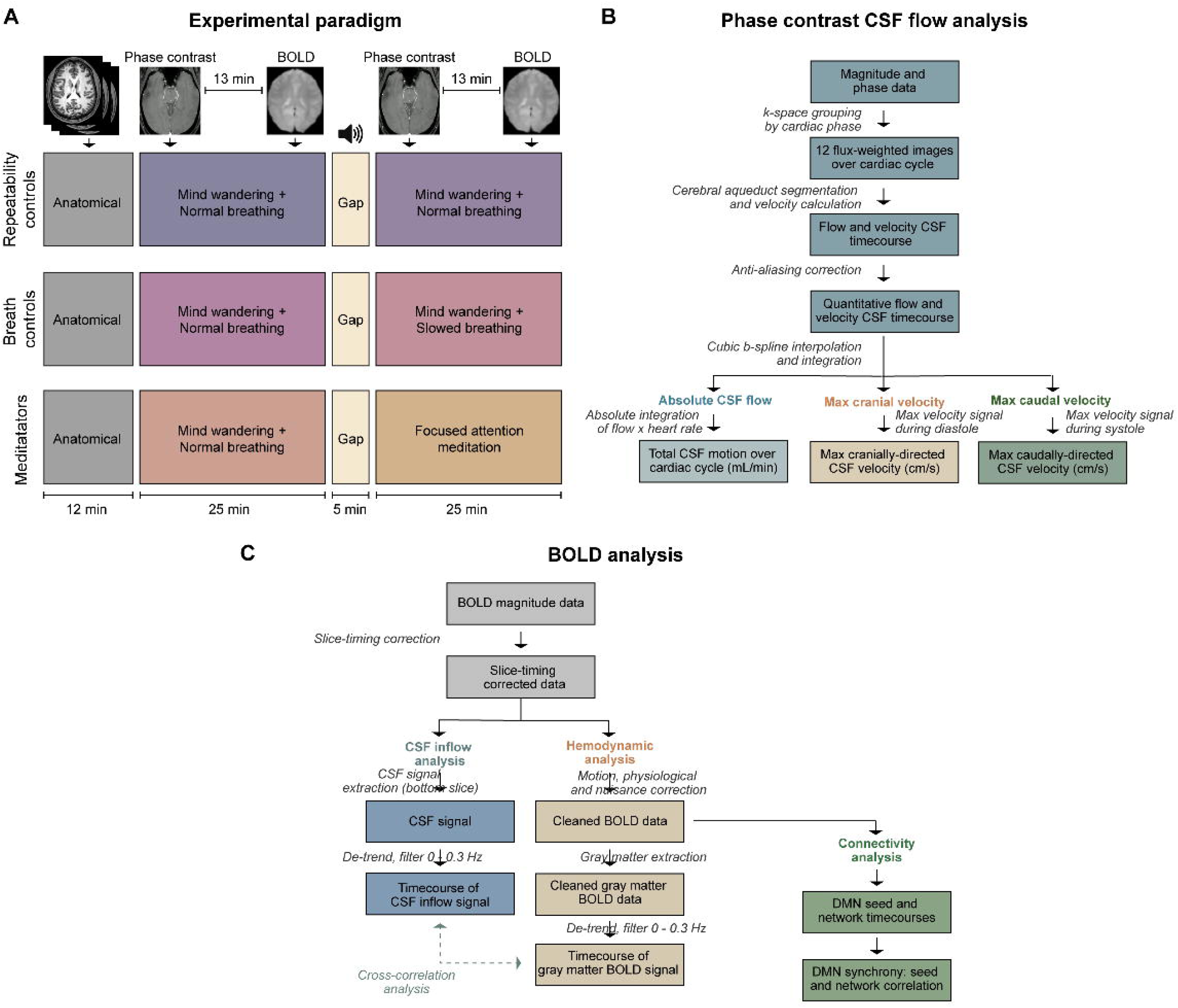
Experimental paradigm and processing overview. (A) The experimental paradigm consists of repeated phase contrast and blood oxygenation level-dependent (BOLD) imaging during mind wandering (MW) and focused attention (FA) meditation, with each epoch spanning approximately 25 minutes and identical timing of each contrast acquisition across participants relative to the FA task. Additional imaging was performed during the 13 min gaps between contrasts, which were not relevant to this study. (B) Overview of the quantitative cerebrospinal fluid (CSF) flow assessments and observables used for hypothesis testing, along with the (C) BOLD analysis pipeline. As the BOLD data were used for (i) CSF inflow analysis, (ii) gray matter hemodynamic assessments, and (iii) default mode network (DMN) connectivity, different processing steps were required for each observable as demonstrated and consistent with prior analogous studies during sleep and meditation. Additional details are provided in **Supporting Information.**

**Table 1.** Summary of demographic characteristics and quantitative cerebrospinal fluid (CSF) flow values for each of the three cohorts. *. Meets stated significance criteria at p < 0.05.

|  | *Repeatability*<br>*controls* |  | *Breath*<br>*controls* |  | *Adept*<br>*meditators* |
| --- | --- | --- | --- | --- | --- |
| **Demographic characteristics** |  |  |  |  |  |
| *Number (count)* | 13 |  | 14 |  | 23 |
| *Age (years)* | 31.9±6.8 |  | 34.6±10.1 |  | 42.3±12.1 |
| *Sex (% female)* | 38.5 |  | 42.9 |  | 39.1 |
| **Respiration (breaths per minute)** |  |  |  |  |  |
| *Scan 1* | 14.6±2.2 | Natural breathing | 14.5±2.1 | Mind wandering | 13.0±2.0 |
| *Scan 2* | 14.6±2.2 | Slowed breathing | 9.8±1.7 | Focused attention | 10.4±2.6 |
| *p-value* | 0.998 |  | <0.001* |  | <0.001* |
| **Absolute CSF flow (mL/min)** |  |  |  |  |  |
| *Scan 1* | 5.22±2.81 | Natural breathing | 4.96±2.05 | Mind wandering | 4.60±2.27 |
| *Scan 2* | 5.26±2.88 | Slowed breathing | 5.01±2.29 | Focused attention | 4.17±2.10 |
| *p-value* | 0.870 |  | 0.845 |  | 0.005* |
| **Max cranial CSF velocity (cm/s)** |  |  |  |  |  |
| *Scan 1* | 3.51±1.36 | Natural breathing | 3.56±1.04 | Mind wandering | 4.26±2.10 |
| *Scan 2* | 3.69±1.45 | Slowed breathing | 3.55±1.13 | Focused attention | 4.03±2.09 |
| *p-value* | 0.085 |  | 0.536 |  | 0.032* |
| **Max caudal CSF velocity (cm/s)** |  |  |  |  |  |
| *Scan 1* | 4.66±1.33 | Natural breathing | 4.77±1.84 | Mind wandering | 4.52±2.14 |
| *Scan 2* | 4.67±1.40 | Slowed breathing | 4.33±1.55 | Focused attention | 4.52±2.23 |
| *p-value* | 0.971 |  | 0.082 |  | 0.985 |

### Confirmation of compliance with meditative task

We assessed task compliance for the meditator cohort using the ordinal self-report scoring (scale=0-10), physiological changes, and the change in DMN network synchrony. On self-report, participants reported a meditation quality of 6.4±1.6 (median=7.0); no participant expressed that they were unable to perform the FA task in the scanner, however, one participant refused to provide a rating. Respiration rate reduced (p<0.001) from 13.0±2.0 (median=13.2) breaths-per-minute during MW baseline to 10.4±2.6 (median=9.3) breaths-per-minute during FA meditation, whereas heart rate reduced (p=0.011) from 73.0±8.2 (median=73.4) beats-per-minute during MW baseline to 67.8±10.6 (median=7.0) beats-per-minute during FA meditation. On functional connectivity analysis, the total explained variance of the DMN was 1.65% during MW baseline and 1.88% during FA meditation, whereas the total variance was 2.97% during MW baseline and 3.28% during FA meditation. Both the increase in total variance (*p*=0.0143) and explained variance (*p*=0.0408) were significant. Functional connectivity, assessed as the cross-correlation coefficient between the standard seed and network region, within the DMN increased (*p*=0.0138) from 0.862±0.090 (median=0.878) to 0.881±0.150 (median=0.926). DMN connectivity analysis findings, which were used only to assist with corroborating the task compliance, including regions assessed, are provided in the **Supporting Information**. Self-report, physiological, and network findings are consistent with the meditators being compliant with the FA task and serve as a prerequisite for subsequent assessments.

### Quantitative CSF flow (cerebral aqueduct)

To test the primary study hypothesis, we applied quantitative phase contrast MRI to investigate whether absolute CSF flow at the level of the cerebral aqueduct differed between MW baseline and FA meditation states, and, whether any observed changes were beyond those assessed during repeated control assessments in MW baseline states with similar changes in respiration rate. **Figure 2** and **Table 1** summarize the absolute CSF flow at the level of the aqueduct assessed from phase contrast MRI. For repeatability controls scanned twice in the same MW state, no change (*p*=0.998) in respiration (14.6±2.2 breaths-per-minute) was observed. Across the cardiac cycle, no significant change was observed in absolute CSF flow (*p*=0.870), maximal retrograde diastolic cranial CSF flow velocity (*p*=0.085), or maximal caudal systolic CSF flow velocity (*p*=0.971) between scans. Next, breath controls with similarly negligible lifetime meditation experience were scanned in sequential identical MW behavioral states, but with instructions to reduce their respiration rate by a target of three breaths-per-minute for scans performed in sequence (i.e., **Figure 1A**). In the first scan, the respiration rate was 14.5±2.1 breaths-per-minute and reduced (*p*<0.001) to 9.8±1.7 breaths-per-minute in the second scan, as intended. Similar to the repeatability scan with no change in respiration rate, no changes in absolute CSF flow (*p*=0.845), maximal retrograde diastolic cranial CSF flow velocity (*p*=0.536), or maximal caudal systolic CSF flow velocity (*p*=0.082) were observed. Finally, in adept meditators scanned first during MW baseline and next during FA meditation (e.g., **Figure 1A**), respiration rate reduced (*p*<0.001) from 13.0±2.0 breaths-per-minute during MW baseline to 10.4±2.6 breaths-per-minute during FA meditation. This paralleled a reduction (*p*=0.005) in the absolute volume of CSF moving across the aqueduct per heart cycle (i.e., absolute CSF flow: total cranially-directed diastolic flow plus total caudally-directed systolic flow) from 4.60±2.27 mL/min during MW baseline to 4.17±2.10 mL/min during FA meditation, which was explained by similar systolic, caudal CSF flow velocities in both states (*p*=0.985) but reduced (*p*=0.032) retrograde cranial CSF flow velocities during FA meditation relative to MW baseline. The above findings are consistent with absolute CSF flow across the cerebral aqueduct reducing during FA meditation due to reduced regurgitant CSF flow during diastole.

**Figure 2.**
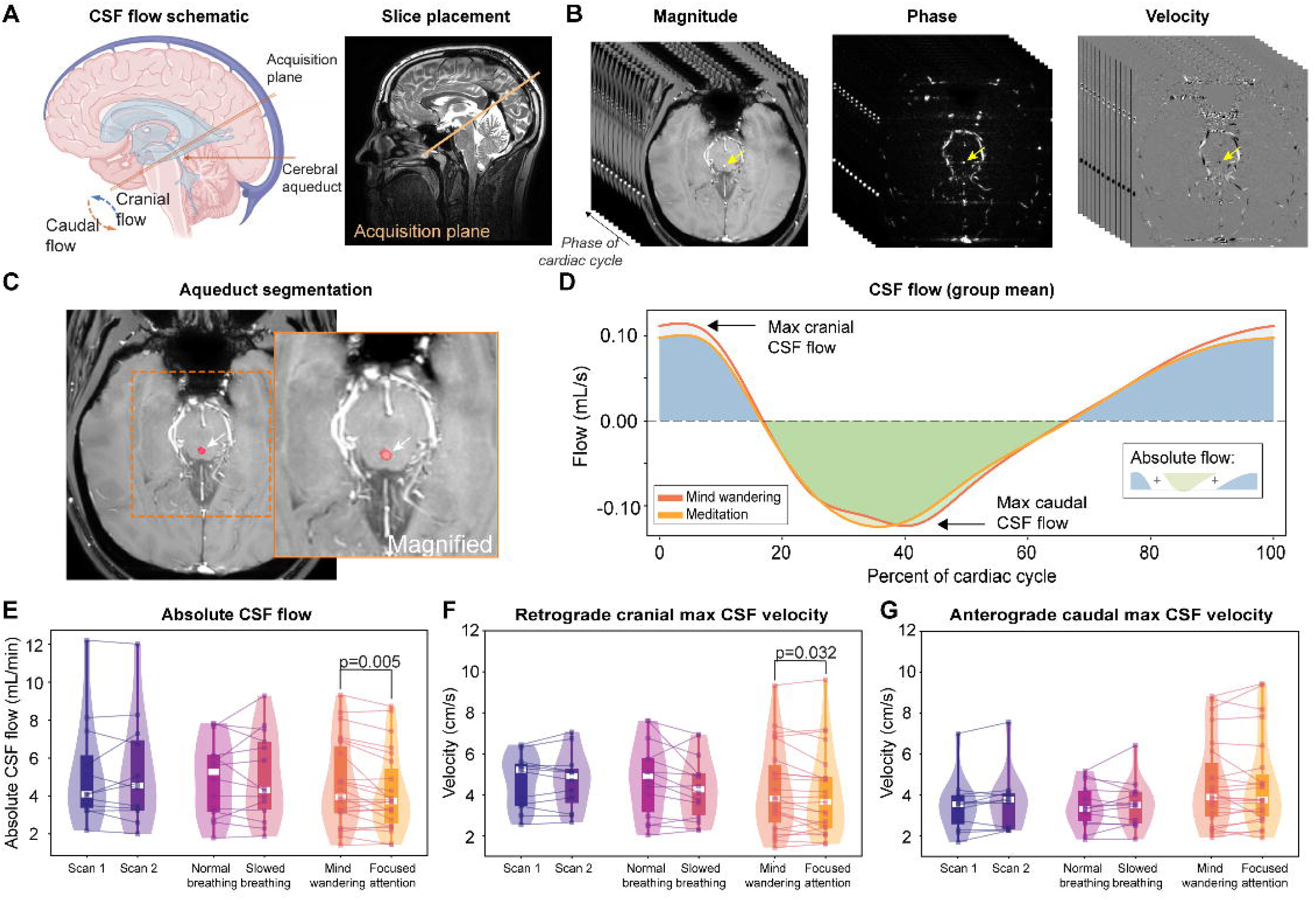
Quantitative cerebrospinal fluid (CSF) assessments during mind wandering (MW) and focused attention (FA) meditation. (A) Schematic of the directionality of CSF flow, along with slice placement on the sagittal 3D *T*_2_-weighted scan. (B) Magnitude and phase data were grouped by cardiac phase to generate CSF flow (mL/s) and velocity (cm/s) time courses (yellow arrow: cerebral aqueduct). (C) Example of the region used for aqueduct (red). (D) Group-averaged (n=23) CSF flow curves for the adept meditators demonstrate that the max diastolic cranially-directed flow is higher in MW baseline compared to FA meditation, and also that the absolute flow (i.e., absolute area under the curve) reduces for FA compared to MW. (E) The heart rate-corrected absolute CSF flow (i.e., absolute flow from (D) multiplied by the heart rate), is similar for repeatability (purple) and breath (pink) controls, but reduces in adept meditators for FA meditation versus MW baseline. This is consistent with (F) a reduction in cranially-directed diastolic flow velocities and (G) unchanging anterograde caudal flow velocities. See **Supporting Information** for additional details on aqueduct segmentation, motion, and quantification considerations.

Finally, baseline states between the control and meditation groups were compared on multiple regression analysis, which revealed that absolute CSF flow in adept meditators was significantly reduced (*p*=0.037; *β*=-1.416) after controlling for age and sex, compared to the control cohort (regression table provided in **Supporting Information**). Supplemental analyses with lifetime meditation experience or self-reported meditation proficiency yielded no trends for significance (*p*>0.500) with CSF flow metrics.

### CSF inflow (cervicomedullary junction)

To test the secondary hypothesis and gain additional information on CSF inflow effects relative gray matter hemodynamic signal fluctuations, similar to what have been investigated during sleep (9), BOLD hemodynamic signal in total supratentorial gray matter and BOLD CSF inflow signal in the bottom slice of the acquisition volume at the approximate level of the cervicomedullary junction were assessed (**Figure 3A-B**). Representative time courses for each region and state are shown in **Figure 3C-D**, whereas group results are summarized in **Figure 3E-F**. The Fourier spectrum of the CSF inflow signal was increased for FA meditation compared to MW baseline, and displayed significant power increases after FDR correction at three frequency bands: 0.0614-0.0887 Hz (*p*=0.018), 0.1535-0.1807 Hz (*p*=0.017), and 0.1814-0.2114 Hz (*p*=0.008). The largest power difference was present at the lower frequency band of 0.0614-0.0887 Hz compared to bands of 0.1535-0.1807 Hz and 0.1814-0.2114 Hz (**Figure 3E-F**). We next evaluated whether the gray matter BOLD signal time course was anti-correlated with the CSF inflow signal, similar to what was reported to occur during sleep (9). No correlation between the time derivative of the gray matter hemodynamic BOLD signal and time derivative of the CSF inflow signal was observed during the MW state (Pearson’s *r*=0.000±0.182; median=-0.022), but an inverse correlation was observed during FA meditation (Pearson’s *r*=-0.091±0.167; median=-0.093), and the change in the correlation coefficient was significant when comparing the MW baseline to FA meditation states (*p*=0.0370). On permutation testing, we observed that the maximal anti-correlation occurred at +0.85s (see **Supporting Information**). These findings are consistent with CSF motion at the approximate level of the skull base increasing, most prominently at the low frequency band of 0.0614-0.0887 Hz, and that the temporal fluctuations in the CSF signal are significantly anti-correlated to the total, supratentorial gray matter hemodynamic fluctuations during FA meditation, but not during MW baseline.

**Figure 3.**
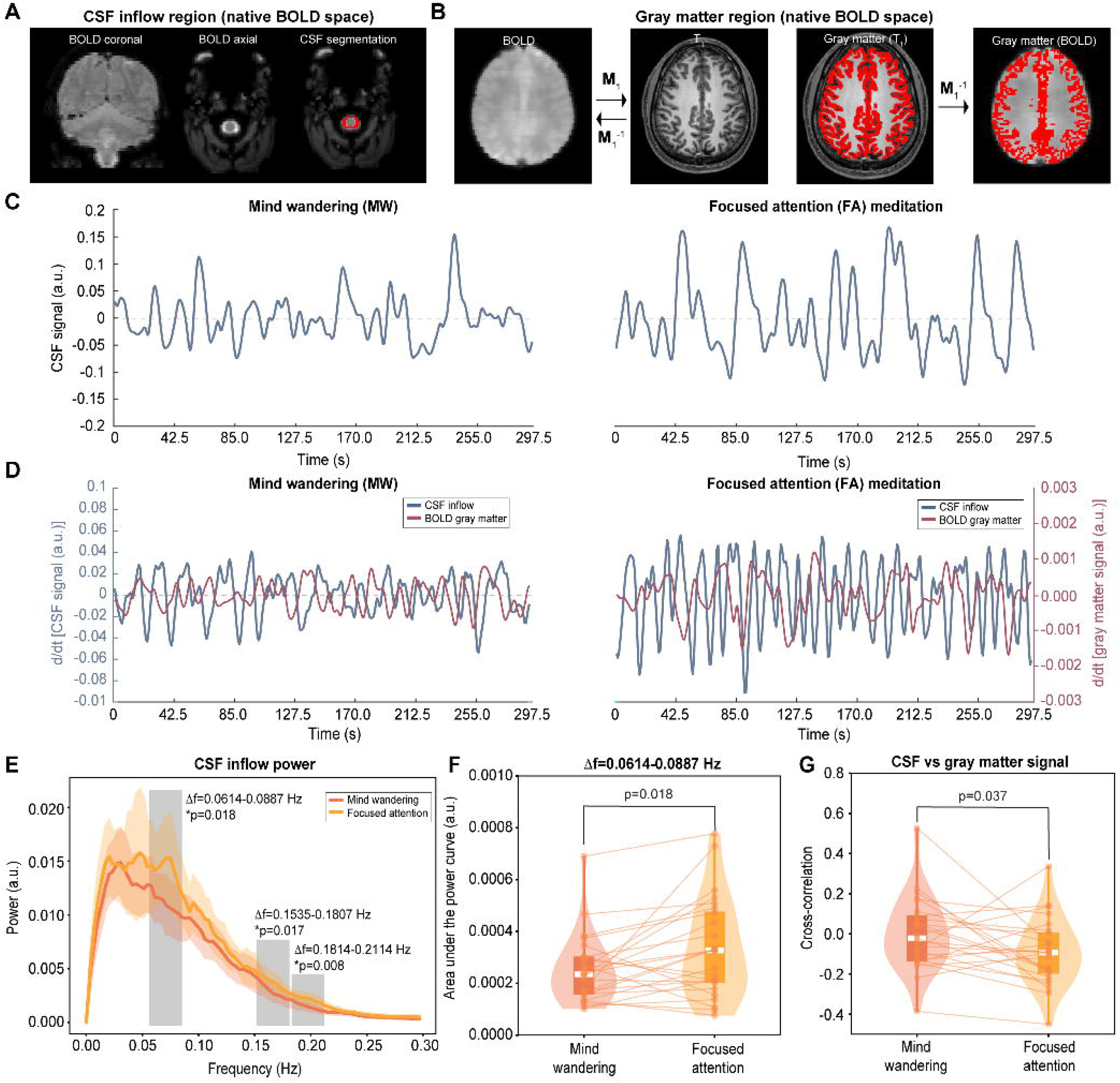
Cerebrospinal fluid (CSF) and hemodynamic assessments during mind wandering (MW) and focused attention (FA) meditation from blood oxygenation level-dependent (BOLD) MRI. (A) Segmentation of the CSF signal in the bottom slice of the BOLD volume, along with (B) example segmentation and registration for gray matter assessment. (C) In a representative participant, the CSF time course shows visible increases in low frequency oscillation transitioning from MW baseline (left) to FA meditation (right). (D) The time derivative of the time courses shows no significant correlation with the time derivative of the gray matter time course for MW baseline, but significant anti-correlation with gray matter during FA meditation. Group results are summarized in (E-F), whereby (E) significant increases in power are observed for FA vs. MW baseline in the CSF time course, with area under the power curve for the highest power of 0.0614-0.0887 Hz curve shown in (F) and anti-correlated CSF and gray matter signal summarized in (G). Different participants are shown in (C) and (D). See **Supporting Information** for additional details on quantification and ancillary permutation testing.

### Motion considerations

Finally, we evaluated whether findings may be attributable to motion-related artifacts, given the change in respiration rate between states. Upon evaluation of the BOLD motion correction parameters, the mean displacement was 0.245±0.135 mm (median=0.18 mm) for MW baseline and 0.264±0.272 mm (median=0.20 mm) for FA meditation, which were not significantly different (*p*=0.705). While the phase contrast data are single slice only, which limit abilities to perform motion correction, we reduced motion sensitivity by focusing on total volumetric flow and max CSF velocity (see **Discussion** and **Supporting Information**), which are both less sensitive to the aqueduct segmentation than other metrics such as mean velocity. Motion parameters and a representative movie of dynamic CSF flow effects are provided in **Supporting Information** to demonstrate motion effects across the two scans. These ancillary analyses provide insufficient evidence to suggest that motion between the two states explain the findings.

## Discussion

Findings demonstrate that a focused attention (FA) style of mindfulness meditation modulates neurofluid dynamics, in the context of previous literature reports (9, 13), in a manner directionally consistent with sleep and opposite to that of aging. Using multimodal MRI methods and accounting for inter-participant heart rate variations, we observed a reduction in absolute CSF flow across the aqueduct during FA meditation, characterized primarily by decreased regurgitant (i.e., cranially-directed) flow during diastole. These reductions were not observed in repeat scans of the same passive, non-meditation mind-wandering (MW) state or when respiration rate was reduced in non-meditators, suggesting that changes in neurofluid flow are not merely artifacts of altered breathing but may reflect state-specific physiological changes induced by meditation. Additionally, we observed that CSF signal fluctuations, assessed near the skull base at the approximate level of the cervicomedullary junction, increased most prominently from MW baseline to FA meditation, within the low frequency band of 0.0614-0.0887 Hz (i.e., 3.7-5.3 cycles per minute). Importantly, while no relationship was observed between such CSF fluctuations and supratentorial gray matter signal fluctuations during MW baseline, FA meditation yielded a significant inverse correlation, suggesting a stronger, inverse coupling between physiological and hemodynamic oscillatory processes. These findings mirror those previously identified during NREM sleep and oppose patterns typically observed with increasing age and neurodegeneration (2, 9, 14). One explanation for these findings is that FA meditation reduces regurgitation of diastolic cranially-directed CSF flow at the level of the aqueduct, improving efficiency and directional coherence of CSF flow. This change parallels increased low frequency signal pulsations in CSF near the skull base, which may reflect improved neurofluid circulation and cerebral waste clearance.

### Physiology of focused attention meditation

As compliance with a FA meditation in a scanner environment is not trivial, we recruited adept meditators with an average 3,708 lifetime hours of meditation experience (median=2,390 hours). On self-report from the ordinal quality of meditation scoring, all participants expressed that they were able to perform the task during the scan. In this cohort, we also observed significant reductions in heart rate and respiration rate during the FA meditation task, along with changes in DMN functional connectivity. These findings are consistent with extant literature (21–25) and the meditators performing the FA task.

The findings can also be considered in the context of what is known regarding the physiology of FA mindfulness meditation. Mindfulness involves a systematic form of mental training that cultivates non-evaluative present-moment awareness by repeatedly regulating attention and arousal while strengthening meta-cognitive processes (27). FA and Open Monitoring (OM) are two complementary styles of core mindfulness practice. FA anchors attention on a single, easily monitored sensory object, typically an external visual target that captures tactile sensations of the breath, and whenever distraction is detected, redirects attention back to that target without judgment (28). OM adopts a receptive, equanimous stance in which attention observes the unfolding stream of sensations, thoughts, and feelings exactly as they arise and pass, without selecting, suppressing, or elaborating on any one event (28). Thus, FA strengthens the capacity to select and sustain a narrow attentional focus, whereas OM cultivates non-reactive awareness across the broad field of experience. Both styles form the foundation of mindfulness-based interventions, now used for a range of physical and mental disorders. The neurobiological mechanisms mediating their health benefits remain incompletely understood (27, 29–31).

Mindfulness has been shown to elicit a constellation of physiological shifts, including lower sympathetic drive, slower cardiorespiratory rhythms, reduced blood pressure and stress hormones, and dampened inflammatory tone, all of which may contribute to its health promoting effects and create a milieu favorable for neurofluid kinetic changes (30). Among mindfulness techniques, the breath-anchored FA meditation has shown to reliably increase parasympathetic dominance, lowering respiratory frequency roughly 15– 30% more than OM, is accompanied by larger vagal heart rate variability gains, and shows greater reductions in salivary cortisol and skin conductance than OM (32). Similar to sleep, such changes have been shown to widen perivascular spaces and ease vascular resistance (33). Experienced mindfulness practitioners have also been found to have unique wake and sleep physiology (17, 34), suggesting a potential neurovascular coupling mechanism that may be conducive for modulating bulk CSF flow or glymphatic activity. Greater fronto-parietal alpha-theta (<12 Hz) and parieto-occipital gamma (25–40 Hz) have been observed during passive rest, during particular styles of meditation, and during NREM stages of sleep (35–37). Changes in DMN functional connectivity during FA have also been observed as a potential biomarker (21–25), and similar changes have been observed here. Theta oscillations drive CSF inflow in the hippocampus (37), and gamma (40 Hz) light-sound stimulation may accelerate glymphatic clearance (38). Together, the converging evidence suggests that mindfulness and particularly FA styles of mindfulness meditation may reveal a plausible, non-pharmacologic route for enhancing neurofluid circulation and, by extension, complement the neurofluid benefits attributed to sleep.

### Meditation, sleep, and neurofluid dynamics

Our findings align with growing evidence suggesting that mindfulness exerts unique effects on brain physiology and health (39, 40). Meta-analyses of mindfulness-based interventions further suggest that cognitive improvements following mindfulness training are most pronounced in adults over 60 years of age (41). Such neuroprotective benefits of mindfulness may partially overlap with the restorative functions of sleep. Both mindfulness and sleep have been shown to reduce pro-inflammatory cytokines, including IL-6, C-reactive protein, and TNF-α, and downregulate genes associated with inflammatory responses (42). Such effects are thought to be mediated by shared mechanisms, such as parasympathetic activation and upregulation of neuroprotective factors such as brain-derived neurotrophic factor (43, 44). Supporting this connection, transcutaneous auricular vagus nerve stimulation, a method of parasympathetic activation, significantly increased net CSF flow in a rodent model of vascular cognitive impairment (45). These findings suggest that mindfulness, through its influence on stress pathways and neurofluid dynamics, may promote brain health through mechanisms similar to those implicated in sleep’s restorative processes.

Regular meditation practice uniquely influences both sleep quality and architecture. Short-term mindfulness training has been shown to significantly improve sleep quality compared to nonspecific active controls (46), likely due to reductions in worry, ruminative thoughts, and mood disturbances that often exacerbate insomnia (47, 48). Interestingly, long-term meditation practitioners report improved subjective sleep quality despite more frequent awakenings and decreased sleep efficiency, possibly reflecting altered perceptions of restfulness or reduced sleep need due to the lower basal metabolic activity during extended meditation practice during retreats (46, 49).

Notably, long-term meditation practitioners also display distinct EEG patterns across states of passive rest, meditation, and sleep, compared to novices (17, 50). For example, increased gamma power in parietal-occipital regions has been observed in advanced meditators and yogis during passive, non-meditative rest (28, 51), often correlating with meditation expertise (26, 51, 52). FA and OM styles of mindfulness are associated with elevated theta and low frequency alpha activity (4–10 Hz) in frontal midline brain regions (17, 26, 50), while Vipassana practice and non-referential compassion meditation increase parietal-occipital and frontoparietal gamma (35–40 Hz) activity, respectively (28, 53, 54). Both Vipassana and non-referential compassion meditation styles emphasize “open-awareness” and “non-duality,” practices characterized by the absence of deliberate noting, reactivity, or evaluation of experiences. During NREM sleep, frontoparietal alpha-theta (<12 Hz) and parietal-occipital gamma power also correlate with meditation expertise (35–37), suggesting a potential interaction between intensive meditation and sleep physiology that may reflect unique neuroprotective benefits.

Specific brainwave patterns implicated in meditation, such as theta and gamma rhythms, are also known to influence neurofluid circulation. For example, theta oscillations in the hippocampus have been shown to promote CSF inflow and facilitate waste clearance in both animal and human models (55). Moreover, stimulation with gamma waves (40 Hz) through light and sound has been shown to enhance neurofluid dynamics and reduce amyloid-beta levels in mice, likely through aquaporin-4 (AQP4) channel hyperpolarization in astrocyte endfeet (38). These findings suggest that meditation may activate glymphatic activity similarly to exogenous stimulation of the awake brain, reinforcing the idea that both sleep and meditation play complementary roles in modulating neurofluid dynamics, promoting brain resilience, and supporting overall well-being.

### Age, neurodegeneration, and neurofluid dynamics

Previous studies using phase-contrast MRI have shown that absolute CSF flow through the cerebral aqueduct (i.e., total motion over the aqueduct per cardiac cycle) increases with age (13). In a cross-sectional study of 77 healthy adults, such higher absolute CSF flow was associated with increasing age (*p*=0.002) (13). Similarly, elevated absolute CSF flow through the aqueduct has been observed in individuals with pre-manifest and motor-manifest Huntington disease (14). These findings suggest that increased total CSF motion through the aqueduct (i.e., cumulative systolic anterograde and diastolic retrograde flow) may serve as clinical biomarkers of declining brain health. In the present study, we found a decrease in absolute CSF flow at the level of the aqueduct when transitioning from a MW baseline to a FA meditative state, directionally consistent with changes that occur in younger rather than older age. Interestingly, this change did not parallel an increase in systolic anterograde flow but rather a similar systolic anterograde flow with a reduced regurgitant retrograde diastolic CSF flow. These findings suggest an increase in the efficiency of CSF flow through the aqueduct with FA meditation.

In our frequency-domain analysis of CSF inflow, from the BOLD data at the approximate level of the skull base, the most robust and significant signal increases occurred within the 0.0614-0.0887 Hz range (∼3.7-5.3 cycles per minute). The findings roughly parallel findings from sleep studies, where increased neurofluid movement was observed at the level of the fourth ventricle (2, 9). Using BOLD MRI to assess CSF inflow at the fourth ventricle, Fultz and colleagues found peak inflow power near 0.05 Hz during stable NREM sleep (9), similar to what was assessed here. CSF inflow fluctuations were also found to be anti-correlated with the cortical parenchymal BOLD signal (9). While our observed frequency range was slightly higher than those observed in Fultz and colleagues, careful inspection of the time courses from Fultz and colleagues will reveal that the ranges largely overlap, albeit with the highest statistical changes being observed in their study at a slightly lower frequency. The overlap in oscillatory patterns supports the hypothesis that FA meditation, like sleep, enhance low frequency CSF pulsations. This is further supported by shared physiological characteristics between sleep and meditation, including slowed respiration and heart rates, as well as the generation of low-frequency brain oscillations like frontal midline theta (4–7 Hz) (17).

### Clinical implications

Understanding the physiological mechanisms underlying mindfulness and its benefits is crucial to informing its increasing application in clinical settings, optimizing intervention strategies, and providing insight into how mindfulness supports mental and physical well-being. The current data show a significant decrease in the regurgitant CSF flow velocities at the level of the cerebral aqueduct during FA meditation, a pattern opposite to that seen in Huntington’s disease patients who exhibit increased velocities and compromised cerebral clearance efficiency (14, 56). The current study findings suggest that FA styles of mindfulness meditation may enhance the brain’s intrinsic waste clearance mechanisms and increase clearance efficiency. These findings have implications for conditions associated with altered neurofluid circulation, such as neurodegenerative diseases and acute neurological conditions. Alzheimer’s disease and traumatic brain injury, for instance, are characterized by the accumulation of cerebral peptides, metabolic waste, or blood products, and impaired CSF flow has been implicated in progression of both conditions (1, 5). If mindfulness can modulate CSF dynamics and promote enhanced glymphatic clearance, it may serve as a non-pharmacological approach to mitigating some effects of these neurodegenerative proteinopathies, although this possibility requires further investigation.

### Limitations and future directions

Our adept meditator cohort comprised 25 recruited participants, 23 of whom met inclusion criteria and had complete data sets. As expected, it was challenging to identify adept meditators who met the high level of lifetime meditation experience even over the multi-year duration of this study. However, these participants were well-characterized with clinical history and neuroimaging and less heterogeneous than what may be expected in larger cohorts with less restrictive inclusion criteria. Nonetheless, future studies with a larger sample size and broader variability in meditative expertise would be useful to further inform these results. Such efforts will require multi-site collaborations to ensure adequate sample diversity and statistical power. In addition, employing both cross-sectional and longitudinal designs can help delineate how specific styles of meditation and duration of practice influence neurofluid dynamics over time. Future research should also examine how different meditation styles affect neurofluid circulation and brain network dynamics. Exploring meditation’s impact on neurofluid dynamics in clinical populations, including individuals with neurodegenerative diseases or sleep disorders, may also identify therapeutic applications. Finally, we included a secondary control cohort with a matched respiration rate change to that of the FA meditation cohort, however, without a wider range of respiration rate modulation it is not possible to fully quantify the impact of respiration rate on several of the measures. As such, future work may also benefit from further investigating how respiration rate vs. FA meditation contribute to the physiological changes observed.

## Conclusion

Our findings provide novel evidence that mindfulness meditation can modulate CSF flow, with implications for both bulk CSF circulation and possibly the neurofluid circuit involving interstitial flow along glymphatic pathways. These results highlight the physiological effects of mindfulness and FA meditation practice, suggesting that the practice may enhance neurofluid circulation and facilitate the clearance of cerebral peptides and waste in a manner analogous to sleep and directionally opposite to aging and neurodegeneration.

## Materials and Methods

### Participants

All participants provided informed consent for this prospective study, which was approved by the Vanderbilt University Medical Center Institutional Review Board. All participants were enrolled between August 2021, and September 2023. For quantitative CSF flow assessments, we enrolled three cohorts with the intent of evaluating (i) intrasession repeatability in the same MW baseline state in meditation naïve healthy adults (*repeatability control*), (ii) the impact of slowing respiratory rate in the same MW state in meditation naïve healthy adults (*breath control*), and (iii) changes between MW baseline and FA meditation in adept meditators (*meditators*). The control cohorts consisted of healthy adults with less than 25 hours of reported lifetime mindfulness practice experience, whereas the meditator cohort consisted of adept FA mindfulness meditation practitioners as defined below.

The inclusion criterion for all cohorts was (1) age 18-65 years inclusive. For meditators, additional criteria were (2) a minimum lifetime formal meditation experience targeted to >500 hours (i.e., >27 minutes/day for 3 years), and (3) self-reported proficiency score >3 (1-5 scale with 5 being most proficient and 1 being little-to-no proficiency) with meditation practices core to mindfulness. Common exclusion criteria for all participants: (1) consumption of stimulants (e.g., coffee and energy drinks) or alcohol within 12 hours of study visit, (2) the following classes of agents known to modify cerebral hemodynamics: benzodiazepines, cholinesterase inhibitors, antipsychotics, opioids, and MAO inhibitors, (3) clinical diagnosis of a major neurological or psychiatric condition (e.g., including but not limited to cognitive impairment, stroke, multiple sclerosis, schizophrenia, bipolar disorder, active alcohol/drug abuse, primary sleep disorder), and (4) observed CSF flow disorder on neuroimaging (e.g., hydrocephalus, perivascular space enlargement beyond that expected for age). Mindfulness experience was reviewed by a neuroscientist with more than 15 years of experience in mindfulness research (DV), clinical and medication history was reviewed by a board-certified neurologist with 14 years of experience (DOC), and anatomical images were reviewed by a board-certified radiologist with 12 years of experience (CDM).

### Meditation protocol

All cohorts underwent the same protocol whereby two states were evaluated in the scanner for 25 minutes each; instructions were explained and participants rehearsed tasks outside the scanner prior to participation (**Figure 1**). Participants also received audio recordings of the scanner noise prior to participation and were requested to practice the FA meditation task during the noises. Anatomical scans were conducted for the first 12 minutes of each scan. Meditators were instructed to enter a passive, non-meditative state of MW for their first 25-minute block, in which participants allowed their mind to freely wander with eyes closed, processing any thought, sensation, or emotion that arose without entering a meditative state or falling asleep. For their second 25-minute block, participants entered an FA meditative state focusing on sensations of the breath around the tip of the nose and upper lip. Using a standardized script (**Supporting Information**), meditators were instructed to notice the sensation of the breath as it moved in and out of the body automatically and effortlessly without manipulating the breath in any way. Meditators were allowed approximately seven minutes prior to CSF flow scanning to enter the FA state. For repeated measures, repeatability controls were instructed to enter a passive, mind-wandering, non-meditative state of rest (MW) for two identical 25-minute-long blocks. Breath controls were instructed to naturally breathe for the first 25-minute block and to voluntarily slow their respiration by a target of three breaths per minute for the second 25-minute block. Respiration was measured using a respiratory bellow and respiration rate (breaths-per-minute) was calculated by dividing the total number of expansion-to-contraction cycles by the scan duration. Heart rate was assessed from a four-electrode ECG recording.

### Neuroimaging acquisition

Neuroimaging assessments were conducted at 3-Tesla (Philips Healthcare, Best, The Netherlands) using phased-array 32-channel reception and dual-transmit body coil radiofrequency transmission. Anatomical imaging consisted of 3D *T*_1_-weighted magnetization-prepared-rapid-gradient-echo imaging (echo time (TE)=3.6 ms, repetition time (TR)=8.2 ms, spatial resolution=1×1×1 mm^3^), 3D *T*_2_-weighted volume isotropic-turbo-spin-echo-acquisition imaging (TE=331 ms, TR=2500 ms, spatial resolution=0.8×0.8×0.8 mm^3^), and 2D diffusion weighted imaging (b-value=1000 s/mm^2^, TE=89 ms, TR=3055 ms, spatial resolution=2.5×2.5×2.5 mm^3^).

For quantitative CSF flow assessments at the level of the cerebral aqueduct, four MR-compatible ECG electrodes were placed on the chest for continuous cardiac monitoring and cardiac phase correction. The imaging slice was planned orthogonal to the cerebral aqueduct and above the fourth ventricle, where the aqueduct is bound by the tectum posteriorly. Phase contrast imaging was used with a protocol identical to that previously published (TR=12.0 ms; TE=7.8 ms; spatial resolution=0.59×0.59×4.0 mm^3^) (13). A 12 cm/s velocity encoding gradient was prescribed, and the mean CSF flow profile was reconstructed for 12 measurements over the cardiac cycle (**Supporting Information**).

To assess CSF inflow at the level of the approximate level of the cervicomedullary junction (junction between the inferior medulla and superior cervical spine), BOLD MRI data (i.e., dynamic *T*_2_*-weighted) were collected twice per participant, once during MW baseline and once during FA meditation. The bottom slice was placed at approximately the level of the cervicomedullary junction approximately 1 cm above the level of the foramen magnum. Scan parameters: TR=850 ms; TE=30 ms; multi-band factor=4; volumes=350; spatial resolution=3×3×3 mm^3^. As the quantitative CSF flow assessments from phase contrast and CSF inflow assessments from BOLD comprised separate hypotheses, we desired to reduce inter-subject variability by keeping the timing of each sequence identical relative to the meditation task; **Figure 1A** and **Supporting Information** provide additional details on timing.

### Meditation analysis

Estimated total lifetime formal meditation experience was determined by daily frequency, consistency of practice, and formal meditation retreat experience using an algorithm based on Hasenkamp and colleagues (57). Participants reported a range of experiences from different meditation traditions (e.g., Buddhist Vipassana, Shamatha, Dzogchen and Mahamudra, Isha Yoga Samyama, and contemporary Insight). Minimum experience included consistent daily practice 4 times/week, 20 minutes/day, for ≥1 year and >1 retreat ≥5 days in length with 6 hours of formal practice/day. A total corrected score was then calculated based on self-reported proficiency rating ≥3 (Likert ordinal 1-5 scale: 1=little to no proficiency and 5=most proficiency).

### Neuroimaging analysis

Imaging was assessed by a board-certified radiologist (CDM) to ensure inclusion criteria were met.

To evaluate CSF flow from the phase contrast scan, the directionality of CSF flow over the cardiac cycle was defined as anterograde (cranial to caudal) or retrograde (caudal to cranial), whereby anterograde flow occurs primarily during systole and retrograde flow primarily during diastole. A mask of the cerebral aqueduct was generated from the magnitude data in the phase-contrast acquisition using a U-Net segmentation approach, which was refined using manual tracing (**Supporting Information**). The mask was applied to the modulus of the phase contrast readout to obtain the acquired phase in the moving CSF over the duration of the cardiac cycle. The 12-point CSF trace was interpolated and using the standard formula for the approximation of flow velocity from phase (58), the maximum cranially (i.e., diastolic) and caudally (i.e., systolic) directed velocities were calculated, along with the absolute CSF flow volume over the cardiac cycle, obtained from the absolute integral of the flux vs. time curve and multiplied by the heart rate to control for possible heart rate variability between participants and states. Note that the absolute CSF flow refers to the total amount of CSF that moves through the aqueduct per unit time. Additional details of the analysis, along with representative images and movies of flow over the cardiac cycle, are provided in **Supporting Information**.

For CSF inflow and hemodynamic assessment, BOLD data were utilized and a pattern analogous to that reported in the literature for similar sleep studies was applied (9). Importantly, given a desired sensitivity to different physiological constructs in gray matter (i.e., hemodynamic activity in the absence of nuisance contributions from motion and CSF pulsatility) and CSF (i.e., inflow specifically sensitive to motion and CSF pulsatility), data were processed differently for each region. A schematic of the processing pipelines, along with representative images and outputs of the relevant steps, are provided in **Figure 1C** and the **Supporting Information**.

For gray matter hemodynamic fluctuations, images were slice-time corrected, motion corrected, and brain-extracted (59–61). An independent component analysis (ICA) with a dimensionality of 30 was applied to the BOLD data and the top 15 nuisance regressors (62), consisting of uncorrelated noise, cardiac pulsatility, and motion, were identified and regressed to create a clean BOLD volume. All analysis occurred in the native BOLD space; however, to obtain an accurate gray matter mask, the gray matter was segmented from the high spatial resolution *T*_1_-weighted images (FSL FAST) (59); the BOLD data were co-registered to the *T*_1_-weighted images and the transformation matrix inverted and applied to the gray matter masks to move the gray matter mask to the native BOLD space (60, 61). The BOLD data were filtered (range=0-0.3 Hz), de-trended, and the gray matter time course calculated in the native BOLD space for each participant and each of the two states. See **Supporting Information** for additional details on each step, along with details of the ancillary default mode network (DMN) evaluation, including imaging examples.

For CSF inflow, nuisance regressors as outlined above were not removed, as many of these provided the desired contrast source. Rather, the BOLD data were slice-time corrected. Next, the CSF was manually delineated from the most inferior BOLD slice, where inflow effects were maximal, and identified as hyperintense signal surrounding the spinal column (**Figure 3**). Images were identically filtered at 0-0.3 Hz, de-trended, and the CSF time course recorded for each participant. The Fourier spectrum for each participant was calculated and grouped into nine bins, with each bin comprising a frequency range of approximately 0.0273 Hz and extending to a maximum frequency of approximately 0.25 Hz. The area under the curve from the power spectra, in each bin, was recorded and preserved for hypothesis testing.

### Statistical analyses

Standard descriptive statistics were calculated for participants by group (i.e., repeatability controls, breath controls, and meditators), including means and standard deviations for continuous variables, as well as counts and percentages for categorical or ordinal variables. Respiration rates were compared using a Wilcoxon signed-rank test to determine if differences between behavioral states were significant. DMN synchrony values and total and explained variance were compared between MW baseline and FA meditation states using Wilcoxon signed-rank tests.

To test the primary study hypothesis, the absolute CSF flow (mL/min) across the cardiac cycle was compared separately between each state for the three cohorts using a Wilcoxon signed-rank test; to assess the source of any hypothesized difference, comparisons were repeated for the maximum anterograde and maximum retrograde velocity. Multiple regression analysis was applied using absolute CSF flow in the MW state as the dependent variable and age, sex, and participant group (i.e., adept meditator vs. control) as exploratory variables to assess whether baseline CSF flow profiles differed between groups.

To evaluate the secondary hypothesis related to CSF signal and gray matter fluctuations from BOLD, Wilcoxon signed-rank tests were employed to the binned powers across all participants for the CSF time courses, and false discovery rate (FDR) was used for comparison correction with required two-sided FDR *p*-value < 0.05. Anticorrelation between individual FA and MW time courses for the total gray matter time course and CSF time course were evaluated by performing a signed-rank analysis of Pearson correlation values between the two regions across the different states. To obtain additional information regarding possible lags between time courses, time regression permutation testing was performed (see **Supporting Information**).

For all statistical assessments, significance criteria were defined as Benjamini–Hochberg FDR-corrected two-sided p<0.05.

## Data availability

Neuroimaging data and analysis scripts will be made available to investigators with current human subjects protections training upon request.

## Acknowledgments

Funding was provided by NIH/NCCIH 5R01AT011456.

