## Supporting Information for "Neurofluid circulation changes during a focused attention style of mindfulness meditation"

**This PDF file includes:**

Supporting text
Figures S1 to S6
Tables S1 to S5
Legend for Movie S1
Supporting Information References

**Other supporting materials for this manuscript include the following:**

Movie S1

**Supporting Information**

*Methods: Meditation script and instructions*

*The below text was read during participants before the mindfulness meditation MRI scans.*

To be read during anatomical scans:
"The entire scanning session will be around one hour. Please try to remain still but comfortable. We will talk to you over the intercom between scans to be sure you are awake and give you instructions to either enter a passive, non-meditative state of mind-wandering or begin breath awareness meditation. We will refer to the passive non-meditative state as *mind wander*. In this state, you can allow your mind to freely wander with eyes closed. Allow your mind to process any thought, sensation, or emotion that arises without falling into a meditative state or falling asleep."

Meditation Scan: To be read during the gap period (see **Figure 1** in manuscript):
"We will now start the meditation scan. Please try to remain still and stay on task. Keep your eyes closed and try not to fall asleep. There will be approximately 20 minutes duration for you to enter and maintain a focused attention meditative state. We ask that you concentrate on the breath in the traditional [Anapanasati] style of breath meditation. Gently rest your awareness around the nostrils of the nose and upper lip as your focus of attention. Simply notice the sensation of the breath as it moves in and out of your body automatically and effortlessly. Don't try to manipulate your breath in any way. Let the breathing go on comfortably and stay with the sensation. The mind will naturally wander away from the sensation of the breath; simply recognize this tendency, and with self-kindness and ease, bring your awareness back to the breath and maintain alert and focused concentration."

Following the above instructions, approximately seven minutes passes prior to the beginning of the quantitative phase contrast CSF flow scan.

*Methods: Phase contrast cerebrospinal fluid (CSF)*

To quantify cerebrospinal fluid (CSF) flow in the cerebral aqueduct, a phase-contrast MRI sequence (velocity encoding = 12 cm/s, spatial resolution = 0.59 × 0.59 × 4.0 mm) was planned orthogonal to the direction of CSF flow at the level of the cerebral aqueduct, below the lateral ventricles, and at the level where the aqueduct is bordered by the tectum posteriorly. Four MRI-compatible ECG leads were placed and cardiac phase correction was applied during reconstruction using the commercially available software on the Philips Elition scanner for *k*-space grouping, with total scan duration between approximately 5 min and 5min30s depending on heart rate. Velocity maps were calculated offline using a standard velocity model for phase contrast (1, 2). To identify the cerebral aqueduct mask on the axial slice, in separate work a 2D U-Net was trained on amplitude images, from 50 individuals with various neurological conditions in which the cerebral aqueduct was manually delineated, to enable automatic segmentation of the cerebral aqueduct. Each mask was subsequently visually inspected and the mask extended if needed, which occurred with oversight from a board-certified neuroradiologist (CDM). The reason for creating a mask that overestimates the size of the aqueduct is because the total volumetric flow was of interest (e.g., mL of CSF per unit time), as well as the peak velocity within the aqueduct. As such, overestimating the aqueduct size does not introduce quantification errors for these metrics (as volumetric flow outside the aqueduct is zero and maximal velocities occur near the center of the aqueduct), whereas underestimation of the mask can lead to underestimation of flow metrics. This approach also reduces sensitivity to motion, as the approach is not as sensitively dependent on the accurate segmentation of the aqueduct as, for example, a mean velocity measure would be, and it is not unreasonable that small motion may occur over the duration of the experiment (considered separately as described below). Following aqueduct detection, each voxel within the region of interest was corrected for potential aliasing effects due to fluid velocities exceeding the velocity encoding threshold. Finally, temporal up-sampling was applied using cubic b-spline interpolation to improve the precision of the curve-derived CSF motion measurements (temporal samples along the cardiac cycle = 100). The most relevant components of this planning and processing are shown in **Figure 2** of the manuscript text, and additional examples, including a single-subject flow curve, are shown in **Figure S1**.

The observables of interest were the absolute CSF flow over the cardiac cycle (i.e., absolute integral of the flow curve over time, which denotes the total amount of CSF moving through the aqueduct cranially during diastole plus the total amount of CSF moving through the aqueduct caudally during systole) and the peak max velocity in the cranial and caudal direction. The absolute CSF flow was multiplied by the heart rate and reported in units of mL CSF / minute. These metrics were selected as they have been shown to have relevance to aging and neurodegeneration in separate studies (1, 3). and net flow (e.g., caudally-directed flow volume minus cranially-direct flow volume) over the cardiac cycle is generally near zero and can be within measurement error of zero in many participants.

*Methods: Blood oxygenation level-dependent (BOLD)*

The processing of BOLD data for secondary hypothesis testing is intended to parallel literature investigations that have reported findings between gray matter hemodynamic fluctuations and CSF inflow during wakefulness and sleep (4). In addition, we extended the analysis to evaluate one core brain network associated with focused attention (FA) styles of mindfulness meditation, the default mode network (DMN). This latter analysis was only to provide additional information to corroborate that the adept meditators were compliant with the task, with the expectation that DMN synchrony would increase during the FA style of mindfulness meditation as we discuss further below. However, the primary study hypothesis focused on the quantitative CSF flow metrics from phase contrast, and the secondary analysis focused on duplicating a previous analysis in gray matter during sleep (4).

Given that each BOLD analysis evaluates different physiological contrasts based on different aspects of the BOLD signal (e.g., gray matter hemodynamics reflecting changes in paramagnetic venous deoxy-hemoglobin vs. CSF inflow characterized by differences in steady-state and equilibrium CSF signal), unique processing of the BOLD data was required for each of these contrast sources. These methods are detailed below and illustrated schematically in **Figure 1** of the manuscript text. All analyses were performed either using scripts from the FMRIB Software Library (FSL) (5) (version 6.0.5.2) or in Matlab (Mathworks, Natick, Massachusetts; R2023a) as outlined below.

1. *For CSF inflow assessments*, we followed pre-processing steps consistent with the literature comparing wakefulness and sleep (4). Magnitude BOLD data were pre-processed with slice time correction, but no spatial smoothing or motion correction were applied. This approach was chosen to preserve sensitivity to small CSF-filled structures near the cervicomedullary junction. As our most inferior slice was placed at the approximate level of the cervicomedullary junction (approximately 1 cm from the foramen magnum), we considered peri-spinal CSF (primary outcome) rather than fourth ventricle flow, which was less sensitive to inflow in our acquisition. The location of the bottom slice is demonstrated in Figure 3 of the manuscript, and here we include the location on the high spatial resolution $T_2$-weighted scan for additional clarity (**Figure S2**). For the peri-spinal CSF, a region-of-interest was manually drawn (software: *fsleyes*) in the native space of the BOLD image on the bottom slice of the $T_2$*-weighted BOLD acquisition. This region was identified as hyperintense relative to surrounding structures, driven by the higher CSF water density, and longer $T_1$ and $T_2^{(*)}$ relaxation times (e.g., **Figure 3** of manuscript). The region was also selected as inflow effects (e.g., motion related differences in equilibrium and steady-state magnetization) will be largest at the bottom of the imaging volume. Importantly, the geometry and acquisition parameters of the mind wandering (MW) and FA meditation BOLD scans were identical, however, the two runs were separated by approximately 13 minutes (e.g., **Figure 1** of manuscript), allowing for potential subject motion between these scans. While motion correction was not performed for the CSF analysis as motion was specifically of interest, we did evaluate the relevance of motion over the experiment to understand whether this could be a confound. To achieve this, affine registration (FSL command: *flirt*) was applied and displacement values recorded (**Table S1**) (6). In identical regions for both the mind wandering and FA meditation states, the BOLD time course was extracted (FSL command: *fslmeants*), normalized, and low-pass filtered (FSL command: *fslmaths -bptf*) at a frequency range of 0-0.3 Hz, mirroring what was previously performed during CSF inflow studies during sleep (4). The final output of this analysis yielded two time courses per participant for the peri-spinal CSF, separately during the MW and FA meditation states.

2. *To calculate the gray matter region of interest*, the $T_1$-weighted scan (magnetization-prepared-rapid-gradient-echo; spatial resolution=1x1x1 mm$^3$) was first pre-processed to reduce the field-of-view to remove

neck and infratentorial regions (FSL command: *robustfov*) and was subsequently segmented using the FMRIB Automated Segmentation Tool (FAST) (7) to reduce bias fields and provide three components (white matter, gray matter, and CSF). The gray matter component was selected by applying a threshold of 0.5 to the gray matter probability map (**Figure S3** shows a gray matter segmentation example). As all BOLD processing occurred in the native space of the BOLD data, the BOLD and $T_1$-weighted data were both brain extracted (FSL command: *bet*) and the BOLD data were co-registered to the $T_1$-weighted data (FSL command: *flirt*) and the affine transformation matrix saved. The inverse of the transformation matrix was then calculated and applied to the gray matter mask to move this mask into the native space of the BOLD data. A binary gray matter mask was saved which corresponded to total supratentorial gray matter within the native BOLD space for each participant. We removed noise components by performing an independent component analysis (ICA) (FSL command: *melodic*; spatial smoothing = 5 mm; slice time correction; motion correction with MCFLIRT; variance normalization; dimensionality estimation=30). From the ICA output, we identified the top 15 nuisance components based on two standard criteria: (i) lack of spatial similarity to known functional networks and (ii) non-physiological spectral profiles, including high frequency noise indicative of cardiac, respiratory, motion, or motion-related artifacts. Representative examples of these non-physiological noise are shown in **Figure S4**. These top 15 regressors were identified separately in the MW and FA meditation states and regressed from the pre-processed data (FSL command: *fsl_regfilt*); as data will be made publicly available upon request, we have reproduced these components here should other investigators seek to duplicate or extend on these analyses (**Table S2**). The time course from the gray matter mask was then used to calculate a mean gray matter time course (FSL command: *fslmeants*) from the filtered and cleaned functional data, which was subsequently normalized and filtered at a low-pass frequency of 0-0.3 Hz to match the frequency range used for CSF inflow analysis described above. The resulting output of this analysis preserved for hypothesis testing was the cleaned mean gray matter time course, corrected for major nuisance regressors.

3. *To evaluate default mode network (DMN)* and compliance with the FA meditation task, we evaluated how the DMN synchrony changed during the FA style of meditation. To achieve this, we identified the DMN using the data pre-processed as in (2); DMN components are listed in **Table S3** along with calculated variance. Here, the DMN was identified by comparing the ICA outputs with the spatial topography of the DMN from the literature (8). We identified the component, percent of explained variance, and percent of total variance within each of the networks, separately for MW and FA meditation states. Examples of identified networks are shown in **Figure S4**. As the size of the DMN will vary between conditions based on statistical thresholds, and we endeavored to evaluate synchrony in the same conserved region, we created a standard seed and network region-of-interest from the literature and 29,671 participants (8) by setting a $z>3.0$ for the region-of-interest and a $z>13$ (total seed voxels = 300 for an approximate seed volume of 37.5 $mm^3$ in the standard 2 mm atlas space). The common seed and region-of-interest are shown in **Figure S5**, along with the DMN from the original cumulative data set. As all processing was performed in the native BOLD space, the regions were moved into the native BOLD space by taking the inverse of the transformation matrix for the BOLD to $T_1$-weighted and $T_1$-weighted to standard space, and concatenating. Finally, the cross-correlation coefficient between the cleaned BOLD data in the seed and network region-of-interest was calculated for each participant to provide an assessment of synchrony. A signed-rank analysis was applied to evaluate whether the cross-correlation statistic increased from MW baseline to FA meditation.

*Methods: Gray matter and CSF cross-correlation*

Correlation of BOLD gray matter and CSF inflow signals were calculated using normalized cross-correlation. First, BOLD and CSF signals were normalized using the z-score method. MW baseline and FA meditation states were analyzed separately, as articulated in the separate processing steps in **Figure 1** of the manuscript, although figures display the results together for comparison. Two different types of analyses were conducted, (a) time-series were used from both BOLD and CSF data from each individual, separately, and (b) the derivative of the BOLD and CSF signal time courses were processed before conducting cross-correlation analysis. Statistics were computed using a permutation testing procedure where BOLD and CSF signal vectors were randomly permuted in an iterative procedure conducted 5,000 times to assess cross-correlation distribution under the null hypothesis. With regards to the analysis using the BOLD and CSF signal time courses, peak negative correlation was observed during meditation with a normalized correlation of -0.05 (95th confidence interval boundary equal to -0.12 and 0.02), with half of the calculated p-values below 0.13. In comparison, during MW baseline, cross-correlation obtained a peak negative correlation coefficient of 0.01 on average (95th confidence interval boundary equal to -0.05 and 0.07). Following the time course analysis, derivatives of the signal were calculated to focus the cross-correlation analysis on the signal changes. For FA meditation, this analysis indicated greater cross-correlation with, on average, a cross-correlation coefficient of -0.09 (95th confidence interval boundary equal to -0.16 and -0.02), with half of the calculated p-values below 0.07. In comparison, during MW, cross-correlation obtained a peak negative correlation coefficient of -0.01 on average (95th confidence interval boundary equal to -0.07 and 0.05). **Figure S6** summarizes the cross-correlation permutation assessments for the MW baseline and FA meditation states. Importantly, we observed a +0.85s shift in maximal correlation between gray matter and CSF time courses.

*Results: CSF flow metrics at baseline between adept meditators and controls*

While this study focused on CSF flow changes during the FA style of meditation, we also evaluated whether there was any evidence to suggest that baseline CSF flow metrics differed in the adept meditator (n=23) and cumulative (n=27) largely meditation naïve control cohorts. To achieve this, we performed regression analyses using the absolute CSF flow as the dependent variable and age, sex, and group (i.e., control=0 or adept meditator=1) as independent variables. The regression findings are summarized in **Table S4**. We observed that absolute CSF flow was lower (p=0.038) in the adept meditator cohort. As absolute CSF flow increases with age and in the presence of neurodegeneration, this finding is largely consistent with meditation exerting positive impacts on brain health. However, it should be noted that additional explanatory variables may be relevant (e.g., medication history, diet, exercise, and education) and our study was not powered to control for these variables. As such, this finding may represent promising preliminary analysis, but larger and more rigorous studies are required to confirm this possibility.

*Discussion: meditation effects on default mode network (DMN) and heart rate*

We utilized self-report, physiological changes (e.g., heart rate and respiration rate), and DMN synchrony to provide corroborating evidence regarding approximate task compliance. On self-report from the ordinal quality of meditation scoring, all participants expressed that they were able to perform the task during the scan. In our cohort, we also observed significant reductions in heart rate (p=0.011) and respiration rate (p<0.001) during the FA meditation task, along with increases in DMN functional connectivity. These findings are consistent with extant literature showing changes in DMN in the setting of mindfulness meditation (9-13) and the meditators performing the FA task.

Across the literature, heart-rate responses during meditation have been reported to be heterogeneous and style-dependent:

a. Breath-anchored, low-cognitive-load practices (e.g., classic relaxation-response mantra or simple breath FA) tend to maintain or slightly lower heart rate, aligning with a parasympathetic, hypometabolic pattern.
b. Practices that add active affective generation or complex attentional monitoring (e.g., slow segmented breathing) can elevate mean HR by ~3–5 bpm, despite subjective calm, suggesting that greater mental effort and sympathetic–vagal co-activation support the task demands.
c. High-frequency respiratory techniques (e.g., breath-of-fire) elicit the largest HR increases (>7 bpm) and reduced HR-variability, reflecting pronounced sympathetic drive.

These observations clarify that an FA style of breath meditation offers the cleanest analogue to the sleep-like, low-HR state relevant for probing neurofluid dynamics, whereas more arousing styles provide a contrasting sympathetic condition useful for mechanistic comparison. **Table S5** summarizes literature on heart rate changes with meditative practices.

*Discussion: Differences between literature and current analysis*

For the secondary analysis of gray matter hemodynamic fluctuations, we attempted to mirror a previously published analysis (4) with the intent of comparing our findings during FA meditation to prior findings during sleep. Our analysis pipeline was extremely similar, however, three differences should be noted. First, the paper by Fultz et al stated that BOLD “raw” data were used, whereas we began our processing with the magnitude data as it was unclear what additional information the raw data would provide for this analysis. Second, while both analyses focused on the bottom slice of the volume to increase sensitivity to inflow effects, our bottom slice was placed at the approximate skull base and level of the cervicomedullary junction, whereas the Fultz et al study placed the bottom slice at the level of the fourth ventricle. Both the fourth ventricle and peri-spinal CSF that we analyzed exhibit bidirectional flow in a manner that is cardiac phase dependent. The low frequency reported pulsations are likely present in both regions, however, it should be noted that different regions were considered. In supplementary analyses we also considered the fourth ventricle, however, given our slice placement we were insufficiently sensitive to inflow effects. Finally, the Fultz et al study filtered their data with a maximum frequency of 0.1 Hz, whereas our filtering extending to 0.3 Hz. This was because we observed statistically significant, albeit low power, differences in CSF time course power near 0.2 Hz, in addition to the very low < 0.1 Hz frequency range. We desired to include this for transparency, however, it should be noted that the greatest power and differences were approximately consistent with the Fultz study and present in the low frequency range below 0.1 Hz.

### Figures

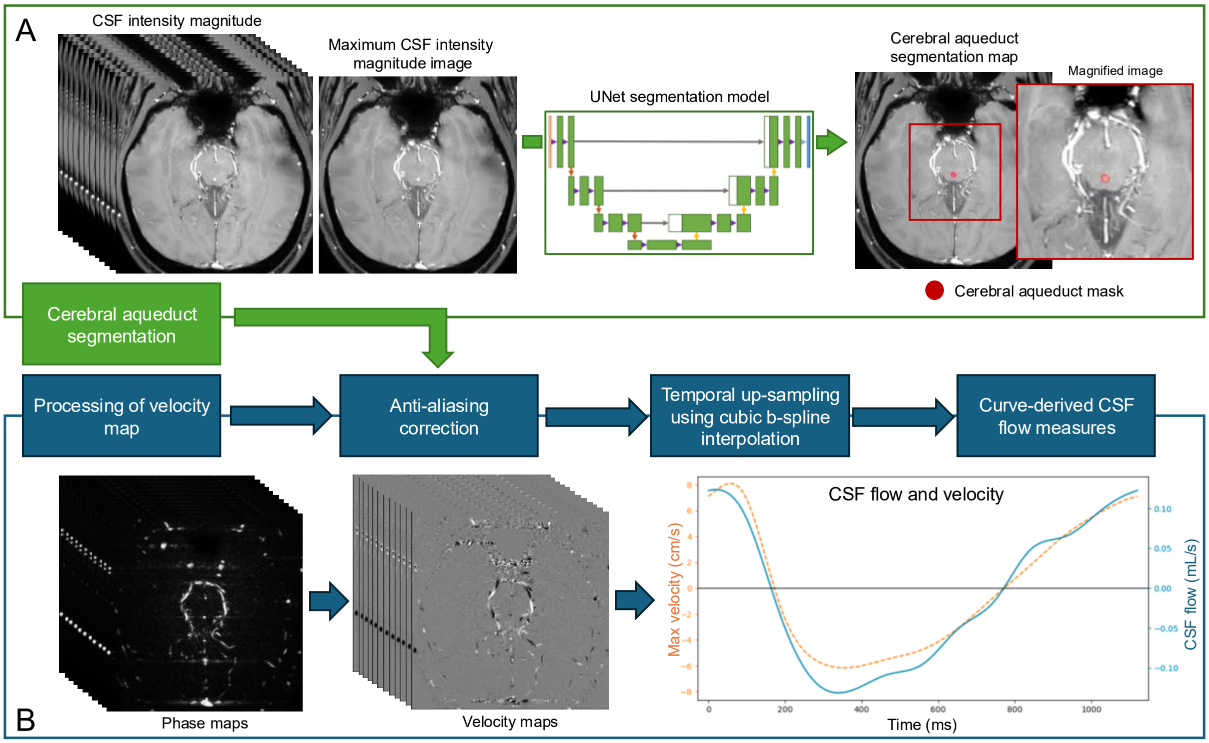


**Figure S1.** Illustration of the pipeline for quantification of the CSF flow using phase contrast magnetic resonance imaging (MRI) planned orthogonal to the direction of CSF flow in the cerebral aqueduct at the level of the tectum. The aqueduct is automatically segmented using a U-Net segmentation model; the mask is subsequently visually inspected and adjusted to overestimate the aqueduct cross-sectional area as needed (see *Methods: phase contrast cerebrospinal fluid).* The curve shown is for a representative single participant, with positive values indicating cranially-directed flow and negative values indicating caudally-directed flow. The group level curve is summarized in **Figure 2** of the manuscript text. Once the absolute flow values are extracted (i.e., total absolute area under the flow curve), this value is multiplied by the heart rate to obtain a final absolute flow value in units of mL CSF / minute.

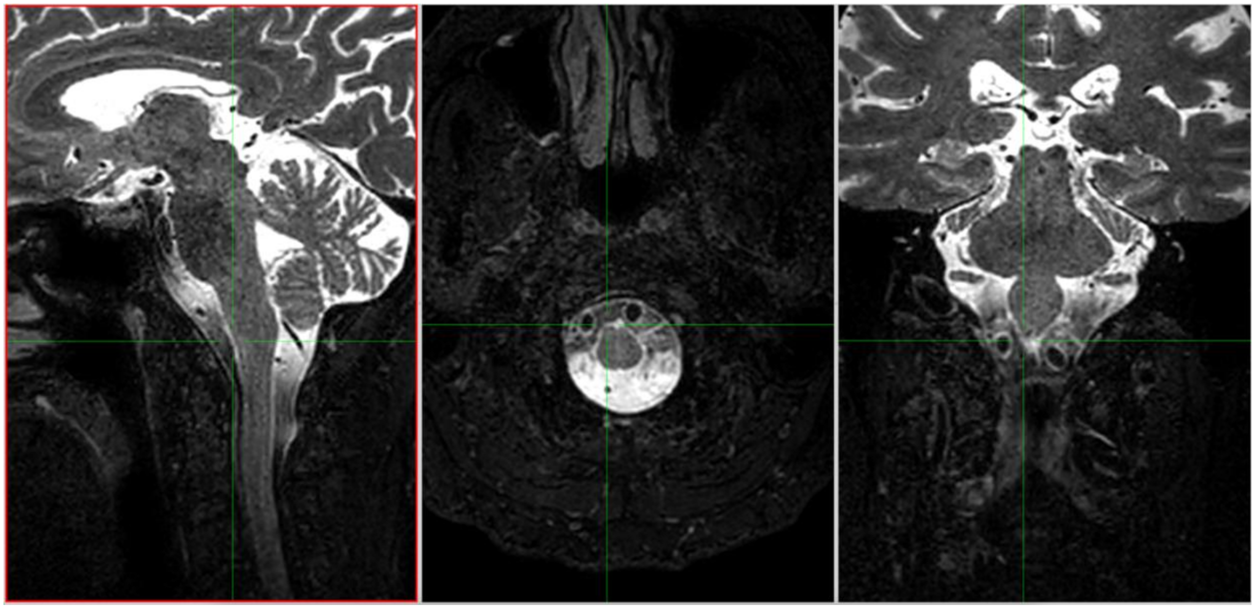

**Figure S2. Location of the bottom slice (e.g., green cross-hairs) for the blood oxygenation level-dependent (BOLD) analysis, shown here on the high spatial resolution $T_2$-weighted scan for clarity.** Cerebrospinal fluid (CSF) inflow was assessed at the approximate level of the skull base and cervicomedullary junction in the CSF surrounding the spinal cord. Sagittal (left), axial (middle), and coronal (right) orthogonal representations are shown; a representative region is also shown in **Figure 3** of the manuscript on the BOLD acquisition itself.

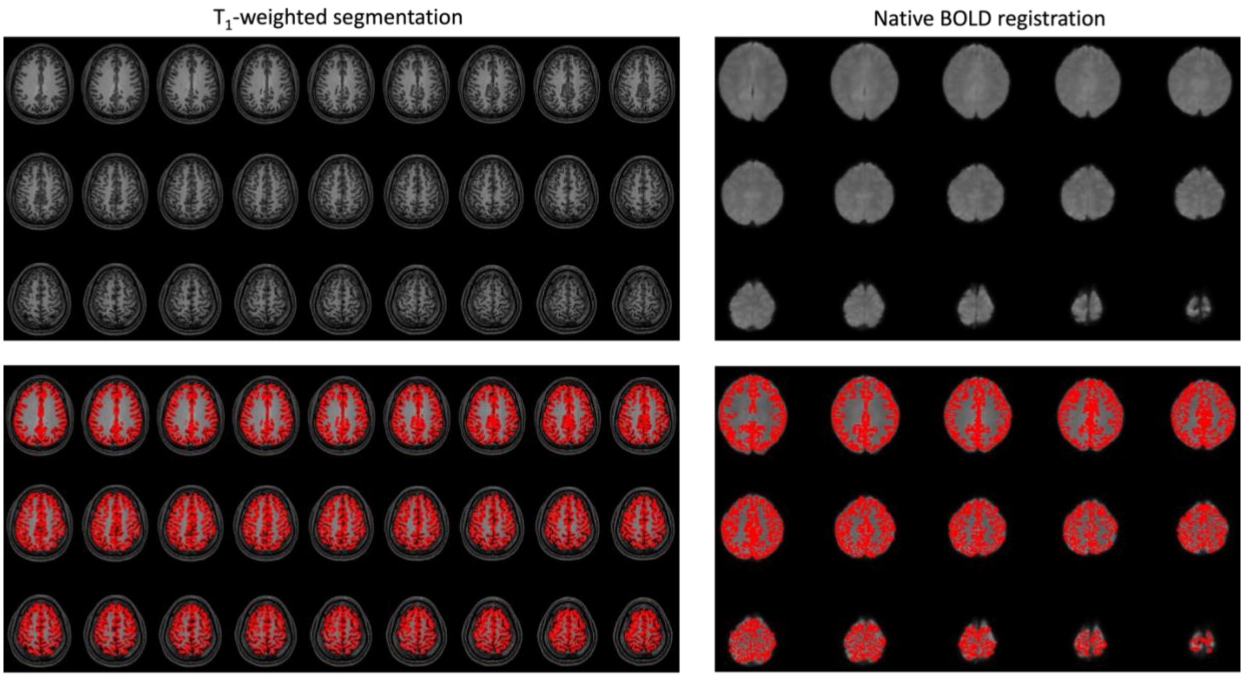


**Figure S3. Gray matter analysis.** Representative slices for total supratentorial gray matter segmentation in native $T_1$-weighted space (left) and back-transformed to the native BOLD space (right). $T_1$-weighted and BOLD data shown above, with overlays of gray matter segmentation (red) shown below. Total supratentorial gray matter was assessed and representative distal slices only are shown.

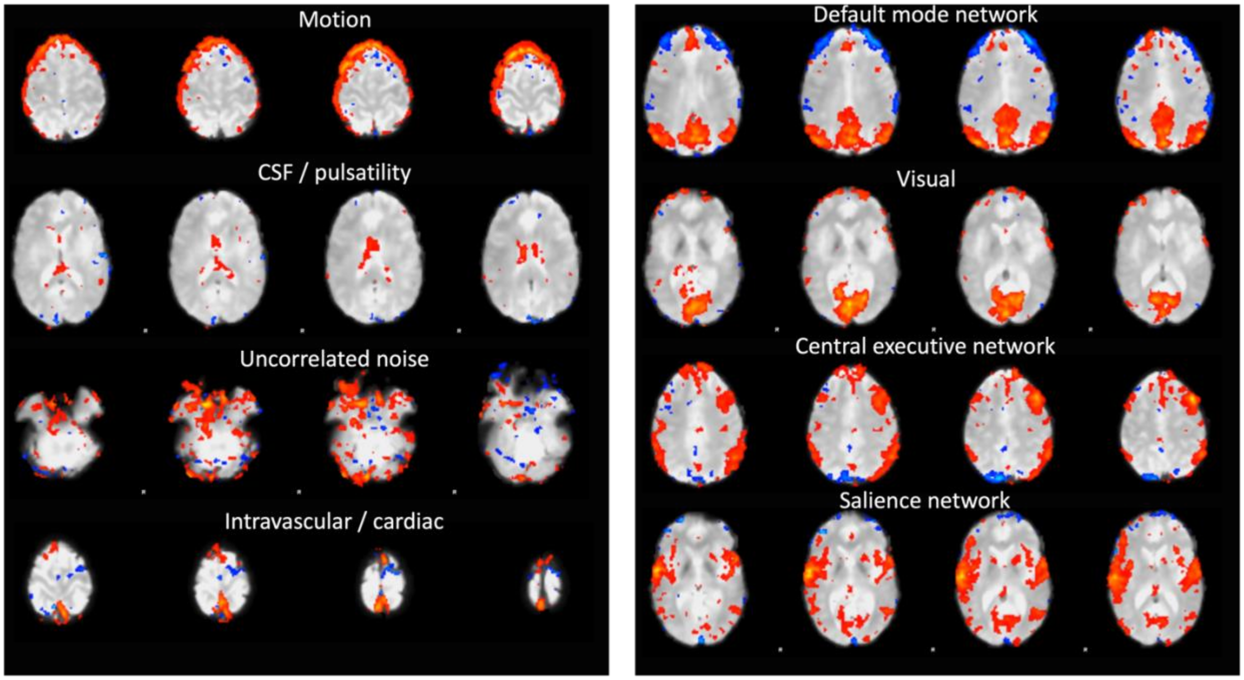


**Figure S4. Representative nuisance regressors (left) and networks (right) identified on independent component analysis (ICA) analysis.** For each participant, the top 15 nuisance regressors were regressed to create a cleaned hemodynamic blood oxygenation level-dependent (BOLD) data set for gray matter time course analysis. Note that the secondary analysis utilized total supratentorial gray matter to parallel prior sleep studies, and this ICA decomposition was used to correct for nuisance contributions in the data.

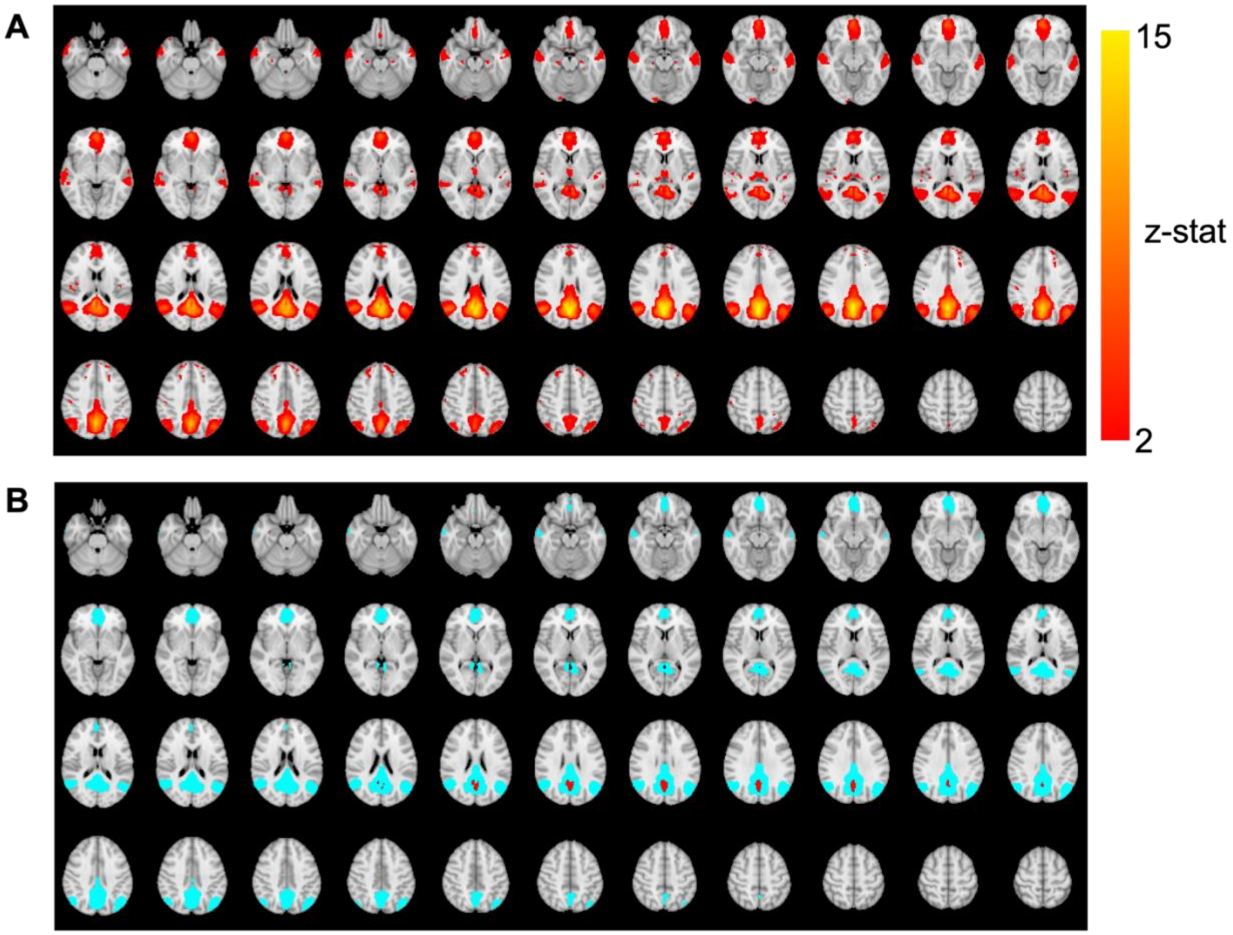


**Figure. S5. Regions used for default mode network (DMN) assessment to corroborate task performance.** (A) The DMN obtained from more than 29,000 data sets (8), along with (B) the thresholded seed (red) and network (blue) region used for synchrony assessments from the cleaned BOLD data. The masks in (B) were transformed to the native BOLD space of each participant. For this analysis, the BOLD images were registered to the $T_1$-weighted image and the $T_1$-weighted image to the standard 2 mm MNI atlas. These matrices were inverted and concatenated to move the standard atlas into the native BOLD space. The above ancillary DMN analysis was only to corroborate task performance, but was not part of testing either the primary or secondary hypothesis.

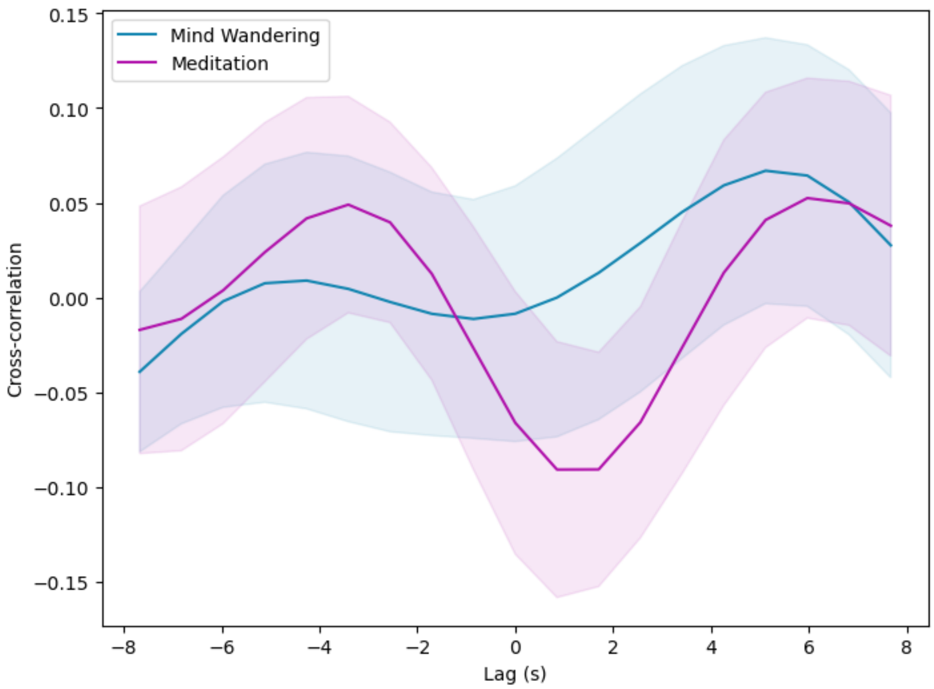


**Figure S6. Cross-correlation of gray matter and cerebrospinal fluid (CSF) blood oxygenation level-dependent (BOLD) time courses.** The blue line denotes the mean cross-correlation value for mind wandering, while purple line denotes the mean of cross-correlation for the focused attention mindfulness meditation. Shaded area represents the 95 percent confidence intervals for each measure across all participants, respectively. Lag corresponds to the delay of BOLD signal in comparison to the CSF signal and is expressed in seconds. The meditation lag reaches a minimum at a lag of +0.85s (i.e., 1 repetition time); during mind wandering, the cross-correlation statistics do not show a clear minimum.

### Tables

| | Mind wandering (MW) | | Focused attention (FA) meditation | |
|---|---|---|---|---|
| Study ID | Mean displacement (mm) | Relative displacement (mm) | Mean displacement (mm) | Relative displacement (mm) |
| *144057* | 0.52 | 0.1 | 0.17 | 0.06 |
| *145017* | 0.39 | 0.08 | 0.17 | 0.05 |
| *145104* | 0.18 | 0.1 | 0.2 | 0.1 |
| *145108* | 0.09 | 0.05 | 0.1 | 0.04 |
| *146897* | 0.16 | 0.09 | 0.28 | 0.06 |
| *146913* | 0.16 | 0.07 | 0.05 | 0.04 |
| *147612* | 0.38 | 0.12 | 0.5 | 0.09 |
| *147904* | 0.16 | 0.08 | 0.16 | 0.08 |
| *151681* | 0.16 | 0.11 | 0.23 | 0.17 |
| *151745* | 0.29 | 0.09 | 0.45 | 0.28 |
| *152087* | 0.46 | 0.07 | 0.2 | 0.07 |
| *153470* | 0.49 | 0.08 | 1.46 | 0.13 |
| *154039* | 0.44 | 0.06 | 0.15 | 0.06 |
| *154118* | 0.14 | 0.05 | 0.1 | 0.05 |
| *157137* | 0.13 | 0.08 | 0.15 | 0.08 |
| *243797* | 0.14 | 0.07 | 0.26 | 0.09 |
| *244044* | 0.36 | 0.08 | 0.25 | 0.05 |
| *244058* | 0.23 | 0.11 | 0.13 | 0.04 |
| *244959* | 0.3 | 0.08 | 0.31 | 0.05 |
| *245911* | 0.24 | 0.06 | 0.21 | 0.03 |
| *249461* | 0.09 | 0.05 | 0.15 | 0.05 |
| *250066* | 0.13 | 0.05 | 0.39 | 0.05 |
| *252055* | 0.1 | 0.11 | 0.13 | 0.1 |
| *252287* | 0.22 | 0.17 | 0.29 | 0.15 |
| *254084* | 0.16 | 0.03 | 0.1 | 0.04 |
| **Mean** | **0.245** | **0.082** | **0.264** | **0.080** |
| **Std dev** | **0.135** | **0.029** | **0.272** | **0.055** |

**Table S1. Motion parameters for BOLD meditation participants.** No significant difference was found for either mean (p=0.705) or relative (p=0.902) displacement between mind wandering and FA meditation. *Note that all 25 recruited participants are included as all data will be made publicly available, whereas two participants were excluded (one lacked phase contrast CSF data and one did not meet radiological inclusion criteria). The statistical significance of this analysis does not change with our without the inclusion of all participants, and as such all acquired data are included for completeness.*

| | Mind wandering (MW) | Focused attention (FA) meditation |
|---|---|---|
| Study ID | Nuisance components | Nuisance components |
| *144057* | 1, 2, 3, 4, 5, 7, 8, 10, 11, 14, 15, 17, 18, 19, 21 | 4, 5, 6, 8, 10, 12, 13, 14, 16, 17, 19, 21, 22, 25, 26 |
| *145017* | 1, 2, 3, 4, 5, 6, 7, 10, 11, 12, 19, 20, 22, 23, 28 | 1, 4, 5, 6, 7, 8, 9, 11, 16, 18, 20, 22, 23, 24, 25 |
| *145104* | 1, 2, 3, 4, 5, 6, 7, 8, 9, 11, 13, 14, 15, 16, 23 | 1, 2, 3, 4, 8, 10, 11, 12, 13, 14, 15, 16, 17, 19, 20 |
| *145108* | 1, 2, 3, 4, 5, 6, 8, 9, 12, 17, 19, 20, 25, 26, 27 | 1, 2, 3, 4, 5, 6, 9, 10, 11, 12, 14, 16, 24, 25, 26 |
| *146897* | 1, 2, 3, 4, 5, 7, 8, 9, 11, 13, 16, 17, 18, 19, 22 | 1, 2, 3, 5, 6, 7, 8, 9, 10, 11, 13, 14, 15, 16, 17 |
| *146913* | 1, 5, 6, 8, 9, 11, 13, 17, 19, 21, 23, 24, 26, 27, 28 | 1, 2, 5, 6, 8, 10, 12, 14, 15, 19, 20, 21, 23, 24, 25 |
| *147612* | 1, 2, 3, 5, 6, 7, 8, 9, 10, 11, 14, 15, 16, 17, 18 | 1, 2, 4, 5, 6, 7, 8, 10, 12, 14, 15, 16, 17, 18, 19 |
| *147904* | 1, 2, 3, 4, 6, 7, 8, 10, 16, 17, 18, 19, 21, 22, 25 | 1, 2, 3, 4, 5, 6, 8, 10, 20, 21, 22, 24, 25, 26, 28 |
| *151681* | 1, 2, 3, 4, 5, 8, 9, 11, 12, 13, 15, 16, 19, 20, 27 | 1, 3, 5, 6, 7, 10, 11, 14, 15, 16, 18, 19, 21, 23, 25 |
| *151745* | 1, 3, 4, 6, 7, 8, 9, 11, 12, 13, 16, 17, 20, 22, 23 | 1, 2, 3, 5, 6, 7, 8, 9, 10, 11, 12, 13, 14, 16, 19 |
| *152087* | 1, 2, 3, 4, 5, 7, 8, 10, 11, 12, 14, 15, 17, 18, 21 | 3, 4, 6, 9, 10, 12, 13, 14, 15, 17, 18, 20, 21, 22, 23 |
| *153470* | 1, 2, 3, 4, 5, 6, 7, 8, 9, 10, 11, 12, 13, 14, 15 | 1, 2, 3, 4, 5, 7, 8, 9, 10, 11, 12, 14, 15, 16, 17 |
| *154039* | 1, 2, 4, 5, 6, 7, 8, 9, 10, 12, 14, 16, 17, 18, 20 | 1, 2, 3, 4, 5, 6, 7, 10, 11, 13, 15, 18, 20, 22, 24 |
| *154118* | 2, 3, 4, 6, 8, 9, 10, 15, 16, 17, 18, 19, 20, 21, 22 | 2, 3, 6, 7, 8, 12, 13, 14, 15, 16, 18, 21, 22, 23, 26 |
| *157137* | 1, 2, 3, 4, 5, 6, 7, 9, 10, 11, 12, 13, 14, 19, 22 | 3, 5, 6, 7, 9, 11, 13, 14, 15, 16, 17, 19, 20, 21, 22 |
| *243797* | 1, 2, 4, 7, 8, 9, 10, 13, 14, 15, 16, 18, 19, 23, 25 | 1, 3, 5, 6, 7, 8, 9, 13, 15, 18, 19, 21, 22, 23, 24 |
| *244044* | 1, 2, 3, 4, 5, 6, 7, 8, 9, 11, 12, 13, 14, 16, 17 | 1, 2, 3, 6, 7, 8, 9, 11, 12, 14, 16, 18, 19, 20, 21 |
| *244058* | 1, 2, 3, 4, 5, 6, 7, 8, 9, 10, 11, 12, 14, 16, 17 | 1, 2, 3, 4, 5, 7, 8, 13, 14, 15, 16, 17, 21, 22, 23 |
| *244959* | 2, 3, 4, 5, 7, 8, 13, 14, 15, 16, 17, 18, 21, 22, 23 | 1, 2, 3, 4, 5, 6, 7, 8, 9, 10, 11, 12, 14, 15, 16 |
| *245911* | 2, 3, 5, 6, 7, 11, 12, 13, 19, 20, 22, 24, 25, 26, 27 | 1, 2, 3, 5, 6, 7, 10, 11, 12, 13, 15, 19, 25, 27, 29 |
| *249461* | 1, 2, 3, 4, 5, 6, 10, 12, 13, 14, 15, 16, 17, 18, 21 | 1, 2, 4, 8, 9, 11, 12, 13, 14, 17, 18, 19, 20, 21, 22 |
| *250066* | 1, 4, 5, 6, 8, 9, 10, 12, 14, 17, 19, 20, 24, 25, 26 | 1, 2, 3, 4, 6, 7, 8, 9, 10, 11, 12, 13, 15, 18, 20 |
| *252055* | 1, 3, 7, 8, 10, 12, 14, 16, 17, 18, 19, 21, 23, 24, 26 | 1, 2, 3, 4, 6, 7, 8, 9, 10, 11, 12, 13, 15, 18, 20 |
| *252287* | 1, 2, 3, 4, 5, 6, 7, 8, 9, 10, 11, 12, 13, 14, 15 | 1, 2, 3, 4, 5, 6, 7, 8, 9, 10, 11, 12, 13, 14, 15 |
| *254084* | 2, 3, 4, 5, 6, 7, 8, 10, 11, 14, 15, 18, 19, 20, 21 | 7, 8, 9, 12, 13, 14, 15, 17, 18, 19, 20, 21, 22, 23, 26 |

**Table S2. Components of the independent component analysis (ICA) 30-component decomposition that were regressed and defined as nuisance contributors.** The list is provided for completeness should other investigators desire to repeat or expand the analysis described here, as BOLD data will be provided upon request. *Note that all 25 recruited participants are included as all data will be made publicly available, whereas two participants were excluded (one lacked phase contrast CSF data and one did not meet radiological inclusion criteria). All participants are included for completeness.*

| | Mind wandering (MW) | | | Focused attention (FA) meditation | | |
|---|---|---|---|---|---|---|
| Study ID | Component | Explained variance (%) | Total variance (%) | Component | Explained variance (%) | Total variance (%) |
| 144057 | 13 | 3.36 | 2.15 | 9 | 3.75 | 1.93 |
| 145017 | 29 | 2.29 | 1.37 | 12 | 3.51 | 2.06 |
| 145104 | 24 | 2.48 | 1.52 | 18 | 3.13 | 1.97 |
| 145108 | 11 | 3.47 | 1.84 | 18 | 3.13 | 1.53 |
| 146897 | 14 | 3.38 | 1.75 | 20 | 2.98 | 1.49 |
| 146913 | 16 | 3.21 | 1.86 | 4 | 4.37 | 1.69 |
| 147612 | 20 | 2.89 | 1.94 | 3 | 4.56 | 3.13 |
| 147904 | 29 | 2.11 | 1.16 | 19 | 2.74 | 1.67 |
| 151681 | 24 | 2.60 | 1.59 | 17 | 2.90 | 1.99 |
| 151745 | 15 | 3.35 | 1.77 | 17 | 3.00 | 2.45 |
| 152087 | 23 | 2.97 | 1.41 | 16 | 3.24 | 1.67 |
| 153470 | 30 | 2.05 | 1.28 | 13 | 3.37 | 2.38 |
| 154039 | 26 | 2.40 | 1.26 | 17 | 3.02 | 1.55 |
| 154118 | 13 | 3.49 | 1.90 | 24 | 2.63 | 1.45 |
| 157137 | 8 | 3.64 | 1.78 | 10 | 3.54 | 1.86 |
| 243797 | 17 | 3.05 | 1.51 | 17 | 2.99 | 1.77 |
| 244044 | 15 | 3.34 | 1.75 | 4 | 4.07 | 2.12 |
| 244058 | 28 | 2.65 | 1.42 | 6 | 3.91 | 1.89 |
| 244959 | 24 | 2.62 | 1.87 | 26 | 2.46 | 1.72 |
| 245911 | 9 | 3.45 | 1.76 | 18 | 3.11 | 1.66 |
| 249461 | 11 | 3.51 | 1.86 | 15 | 2.94 | 1.67 |
| 250066 | 7 | 3.96 | 1.88 | 15 | 3.16 | 1.97 |
| 252055 | 20 | 2.94 | 1.82 | 19 | 3.09 | 1.68 |
| 252287 | 28 | 1.98 | 1.34 | 27 | 2.31 | 1.49 |
| 254084 | 12 | 3.19 | 1.46 | 5 | 4.06 | 2.29 |
| **Mean** | **18.64** | **2.97** | **1.65** | **14.76** | **3.28** | **1.88** |
| **Std Dev** | **7.38** | **0.55** | **0.26** | **6.73** | **0.57** | **0.38** |

**Table S3. Default mode network (DMN) components and explained variance.** *Note that all 25 recruited participants are included as all data will be made publicly available, whereas two participants were excluded (one lacked phase contrast CSF data and one did not meet radiological inclusion criteria). The statistical significance of this analysis does not change with our without the inclusion of all participants, and as such all acquired data are included for completeness.*

| | $\beta$ | *Standard Error* | *t Stat* | *P-value* | *Lower 95%* | *Upper 95%* |
|---|---|---|---|---|---|---|
| Intercept | 1.08 | 1.21 | 0.90 | 0.38 | -1.35 | 3.52 |
| Age (years) | 0.010 | 0.03 | 3.22 | 0.002 | 0.04 | 0.16 |
| Sex (female=0; male=1) | 1.34 | 0.62 | 2.15 | 0.036 | 0.09 | 2.60 |
| Group (control=0; meditator=1) | -1.42 | 0.66 | -2.14 | 0.038 | -2.75 | -0.08 |

**Table S4. Regression results for mind wandering (MW) cerebrospinal fluid (CSF) absolute flow (mL/min).** The findings suggest that after controlling for age and sex, absolute CSF flow is reduced (p=0.038) in the adept meditator (n=23) compared to the cumulative control (n=27) cohort.

| Study | Meditation style(s) and sample | Main HR finding(s) |
|---|---|---|
| Benson et al., 1974 (classic "relaxation-response") | 36 experienced Transcendental-Meditation practitioners | Mean HR reduced by ~3 beats·min$^{-1}$ relative to quiet rest, accompanying the well-known hypometabolic profile |
| Peng et al., 2004 | 10 Kundalini-yoga practitioners performed three 10-min protocols: relaxation-response mantra (RR), rapid "breath-of-fire" (BF), and slow segmented breathing (SB) | *Versus immediate baseline:* RR produced no significant change in mean HR; SB raised HR by ≈ 4 beats·min$^{-1}$ ($p < 0.05$); BF produced the largest rise (≈ 7.6 beats·min$^{-1}$, $p < 0.01$) |
| Lumma et al., 2015 (ReSource Project, 3-month modules) | 157 meditation-naïve adults trained Breathing (FA), Loving-Kindness (LKM) & Observing-Thoughts (OT) | Baseline (week 3): HR ~71 beats·min$^{-1}$ across styles. Training effects (week 13 minus week 3): HR unchanged during Breathing (+0.8 bpm, n.s.); LKM +4.7 bpmand OT +2.8 bpm (both $p < 0.001$) indicating sympathetic recruitment in these cognitively/affectively demanding practices |

**Table S5. Literature findings on heart rate changes during different meditation styles.** References for Benson et al. (14), Peng et al. (15), and Lumma et al. (16) are included in the supplementary references of this document.

**Movie legend**

**Movie S1 (separate file).** Movie showing the cerebrospinal fluid (CSF) flow signal and level of motion for a representative subject for mind wandering (MW) baseline and focused attention (FA) meditation. Quantitative motion metrics from BOLD scans are provided in **Table S1**.

**Supporting Information References**


1. J. J. Eisma *et al.*, Choroid plexus perfusion and bulk cerebrospinal fluid flow across the adult lifespan. *J Cereb Blood Flow Metab* **43**, 269-280 (2023).
2. E. M. Haacke, *Magnetic resonance imaging : physical principles and sequence design* (Wiley, New York, 1999), pp. xxvii, 914 p.
3. K. Hett *et al.*, Cerebrospinal Fluid Flow in Patients with Huntington's Disease. *Ann Neurol* **94**, 885-894 (2023).
4. N. E. Fultz *et al.*, Coupled electrophysiological, hemodynamic, and cerebrospinal fluid oscillations in human sleep. *Science* **366**, 628-631 (2019).
5. M. Jenkinson, C. F. Beckmann, T. E. Behrens, M. W. Woolrich, S. M. Smith, Fsl. *Neuroimage* **62**, 782-790 (2012).
6. M. Jenkinson, P. Bannister, M. Brady, S. Smith, Improved optimization for the robust and accurate linear registration and motion correction of brain images. *Neuroimage* **17**, 825-841 (2002).
7. Y. Zhang, M. Brady, S. Smith, Segmentation of brain MR images through a hidden Markov random field model and the expectation-maximization algorithm. *IEEE Trans Med Imaging* **20**, 45-57 (2001).
8. S. M. Smith *et al.*, Correspondence of the brain's functional architecture during activation and rest. *Proc Natl Acad Sci U S A* **106**, 13040-13045 (2009).
9. C. C. C. Bauer, S. Whitfield-Gabrieli, J. L. Diaz, E. H. Pasaye, F. A. Barrios, From State-to-Trait Meditation: Reconfiguration of Central Executive and Default Mode Networks. *eNeuro* **6** (2019).
10. I. Sezer, D. A. Pizzagalli, M. D. Sacchet, Resting-state fMRI functional connectivity and mindfulness in clinical and non-clinical contexts: A review and synthesis. *Neurosci Biobehav Rev* **135**, 104583 (2022).
11. A. Berkovich-Ohana, M. Harel, A. Hahamy, A. Arieli, R. Malach, Alterations in task-induced activity and resting-state fluctuations in visual and DMN areas revealed in long-term meditators. *Neuroimage* **135**, 125-134 (2016).
12. V. A. Taylor *et al.*, Impact of meditation training on the default mode network during a restful state. *Soc Cogn Affect Neurosci* **8**, 4-14 (2013).
13. J. A. Brewer *et al.*, Meditation experience is associated with differences in default mode network activity and connectivity. *Proc Natl Acad Sci U S A* **108**, 20254-20259 (2011).
14. H. Benson, B. A. Rosner, B. R. Marzetta, H. P. Klemchuk, Decreased blood pressure in borderline hypertensive subjects who practiced meditation. *J Chronic Dis* **27**, 163-169 (1974).
15. C. K. Peng *et al.*, Heart rate dynamics during three forms of meditation. *Int J Cardiol* **95**, 19-27 (2004).
16. A. L. Lumma, B. E. Kok, T. Singer, Is meditation always relaxing? Investigating heart rate, heart rate variability, experienced effort and likeability during training of three types of meditation. *Int J Psychophysiol* **97**, 38-45 (2015).
